# Public awareness of and opinions on the use of mathematical transmission modelling to inform public health policy in the United Kingdom

**DOI:** 10.1101/2023.07.31.23293324

**Authors:** Ruth McCabe, Christl A. Donnelly

**Affiliations:** Department of Statistics, University of Oxford; NIHR Health Protection Research Unit in Emerging and Zoonotic Infections; Pandemic Sciences Institute, University of Oxford; MRC Centre for Global Infectious Disease Analysis, Imperial College London

## Abstract

Mathematical transmission modelling is a key component of scientific evidence used to inform public health policy and became particularly prominent during the COVID-19 pandemic. As key stakeholders, it is vital that the public perception of this set of tools is better understood. To complement a previously published article on the science-policy interface by the authors of this study, novel data were collected via responses to a survey via two methods: via an online panel (“representative” sample) and via social media (“non-probability” sample). Many identical questions were asked separately for the period “prior to” compared to “during” the COVID-19 pandemic.

All respondents were increasingly aware of the use of modelling in informing policy during the pandemic, with significantly higher levels of awareness among social media respondents than online panel respondents. Awareness generally stemmed from the news media and social media during the pandemic. Transmission modelling informing public health policy was perceived as more reliable during the pandemic compared to the pre-pandemic period in both samples, with awareness being positively associated with reliability within both samples and time points, except for social media during the pandemic. Trust in government public health advice remained high across samples and time periods overall but was lower in the period of the pandemic compared to the pre-pandemic period. The decay in trust was notably greater among social media respondents. Many respondents from both samples explicitly made the distinction that their trust was reserved for “scientists” and not “politicians”. Almost all respondents, regardless of sample, believed governments have responsibility for the communication of modelling to the public.

These results provide an important reminder of the potentially skewed conclusions that could be drawn from non-representative samples.

## Introduction

Scientific advisory mechanisms in the United Kingdom of Great Britain and Northern Ireland (UK) were brought into the full view of the public eye during the COVID-19 pandemic. Restrictive physical distancing measures, such as lockdowns, affected all within society and, rightfully, sparked a widespread interest in the evidence underpinning such decision-making by the government. As society reflects on the events of the COVID-19 pandemic, particularly in anticipation of the outcome of the public inquiry into the UK government’s handling of the pandemic,^1^ the use, or not, of scientific evidence to inform policy will be examined closely.

Mathematical transmission modelling is a key component of scientific evidence presented to the government during outbreaks of infectious diseases: the COVID-19 pandemic was no exception.^2, 3^ Already, there is a growing body of work focusing on the use of this set of tools during the recent pandemic,^4–6^ including one by the authors of this study.^2^ In our previously published study, we examined the interplay between mathematical transmission modelling, scientists, the media and the public throughout the first year of the pandemic.^2^ Participants in our research, namely scientific advisors and public communication experts, indicated that direct communication with the public had been challenging. As the success of public health policy often depends upon the cooperation of the public, it is important that the public perception of scientific advice, in this case specifically mathematical modelling, is understood. Much of the research into public opinions throughout the COVID-19 pandemic has related to vaccine hesitancy^7–9^ or adherence to public health and social measures^10–13^. Although Marshall et al.^14^ interviewed members of the public regarding Test, Trace and Isolate policies with a view to improving assumptions underpinning mathematical modelling, respondents were not explicitly asked about their perceptions of modelling.

We sought to complement our initial study^2^ by gathering and analysing data on the public’s awareness of and opinions on the use of mathematical transmission modelling to inform public health policy in the UK via an online questionnaire. Even with sufficient financial resources, obtaining a “representative” sample of the population is notoriously difficult and thus the interpretation of the resulting data must be cautious regarding the opinions of the adult population in the UK. Consequently, we collected data via two sampling methods: via an online panel (using the platform *Prolific Academic*^15^ and considered a “representative” sample) and via social media (using *Twitter*^16^ and considered a “non-probability” sample). As such, the goals of this study are two-fold: to better understand public awareness of and opinions on the use of modelling to inform public health policy but also, critically, to explore and interpret any systematic differences between the two samples obtained.

## Materials and Methods

### Survey design and distribution

This study has approval from the Medical Sciences Interdivisional Research Ethics Committee at the University of Oxford with Ethics Approval Reference R76166/RE001. The Participant Information Sheet (PIS), provided in the Supplementary Material, sets out the instructions given to participants ahead of taking the survey.

We designed an online survey of 24 questions, comprising of 16 multiple-choice and 8 open-ended questions (Table 1). The survey was split into four sections of unequal length.

**Table 1:**
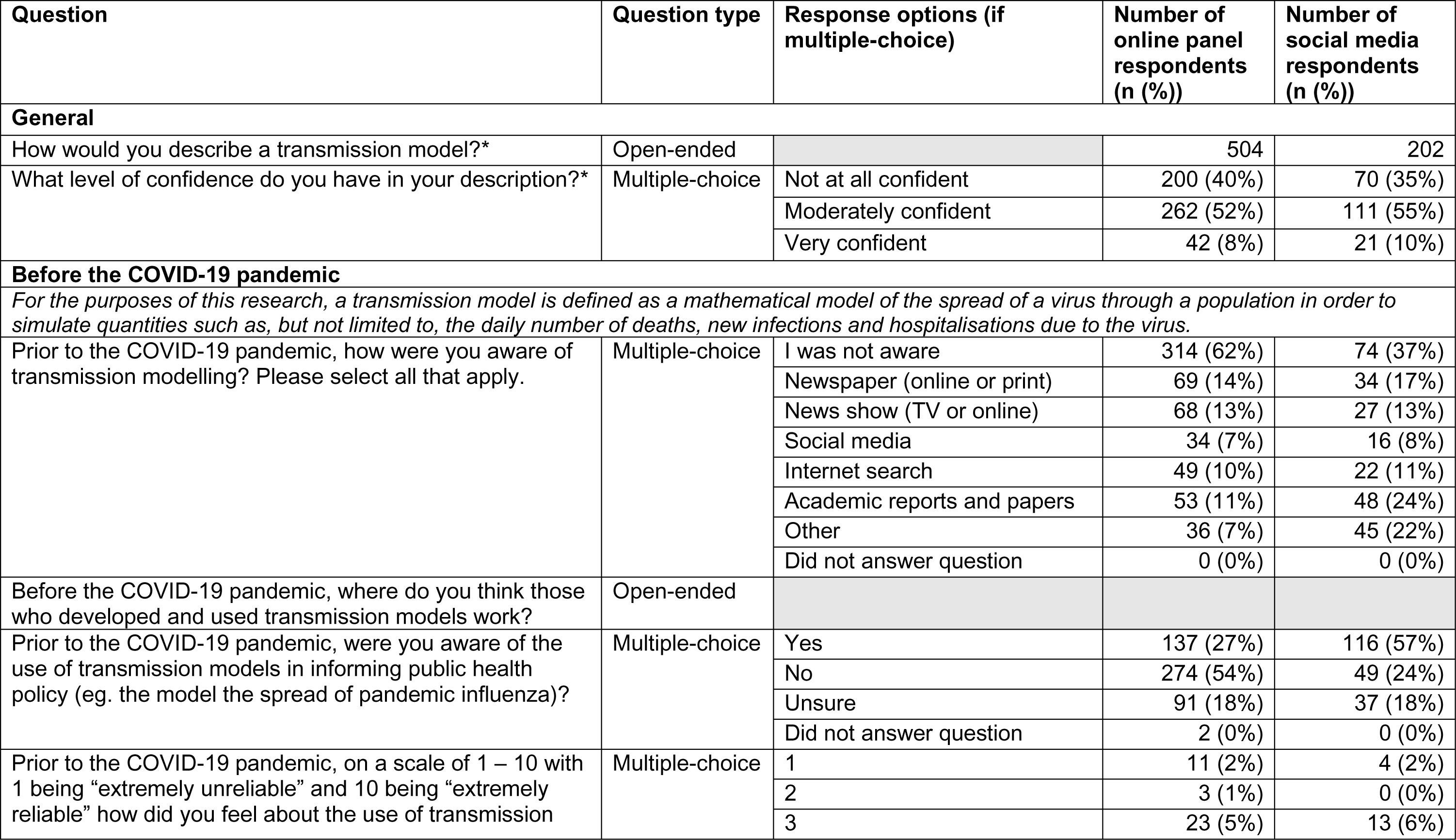
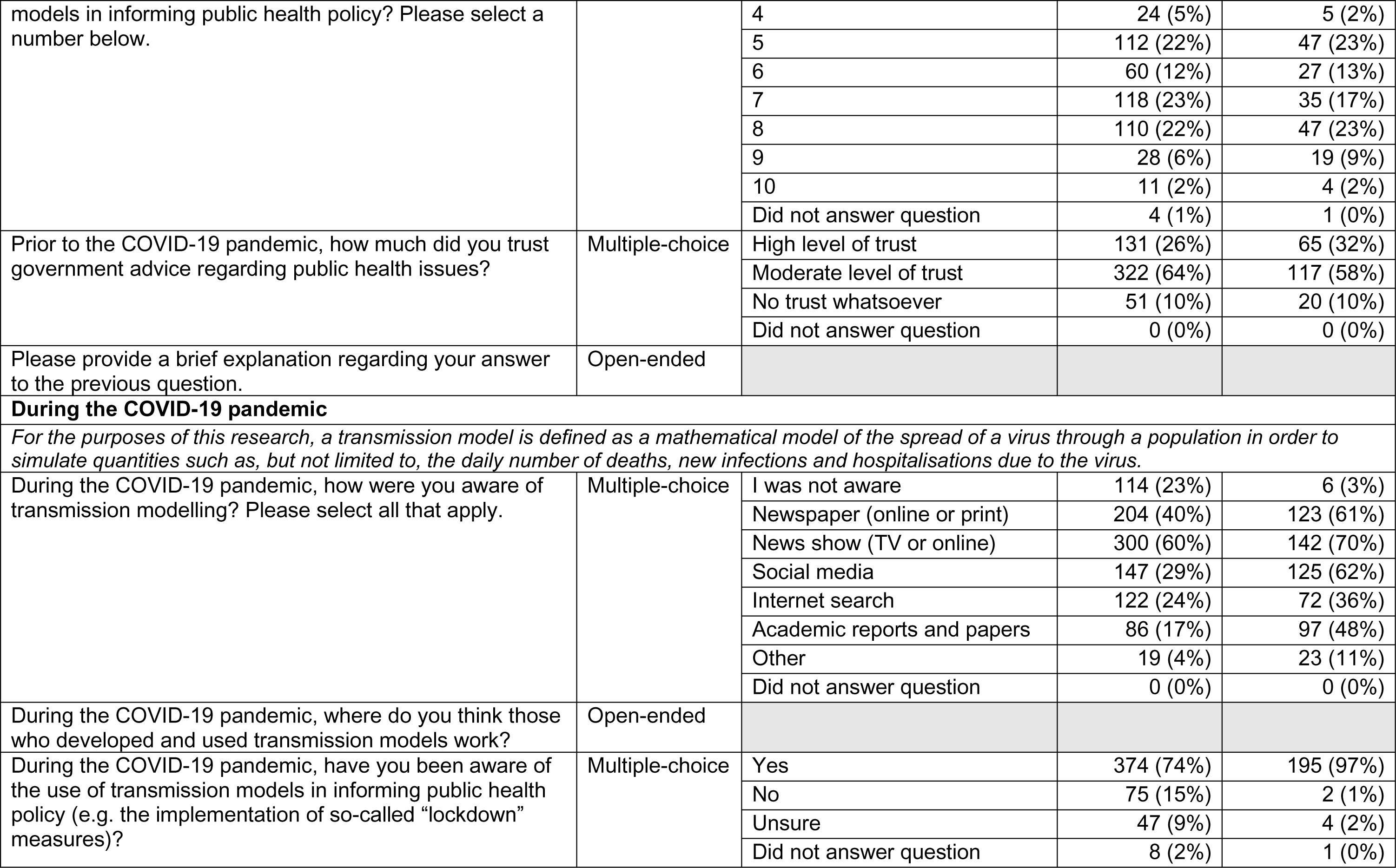
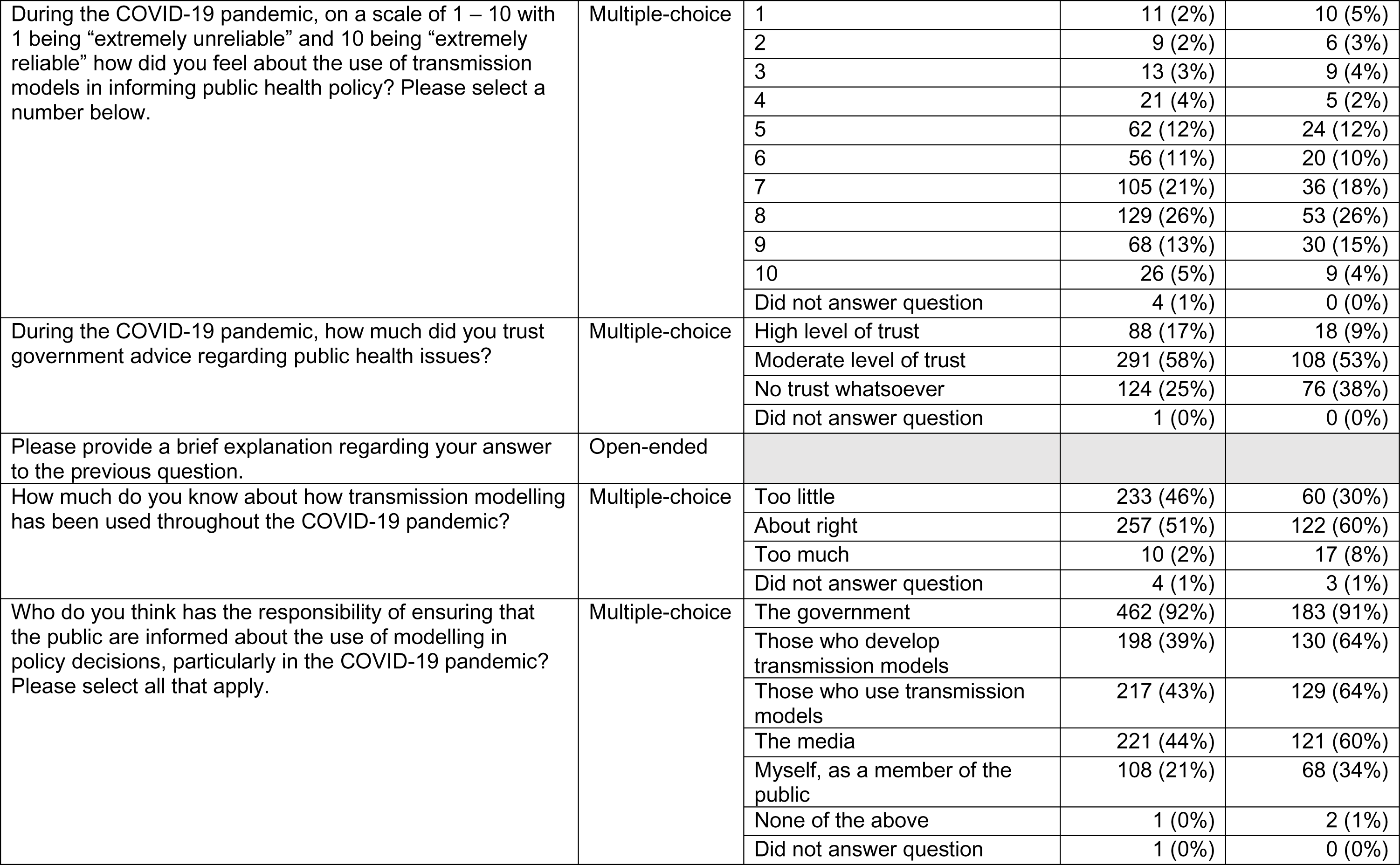
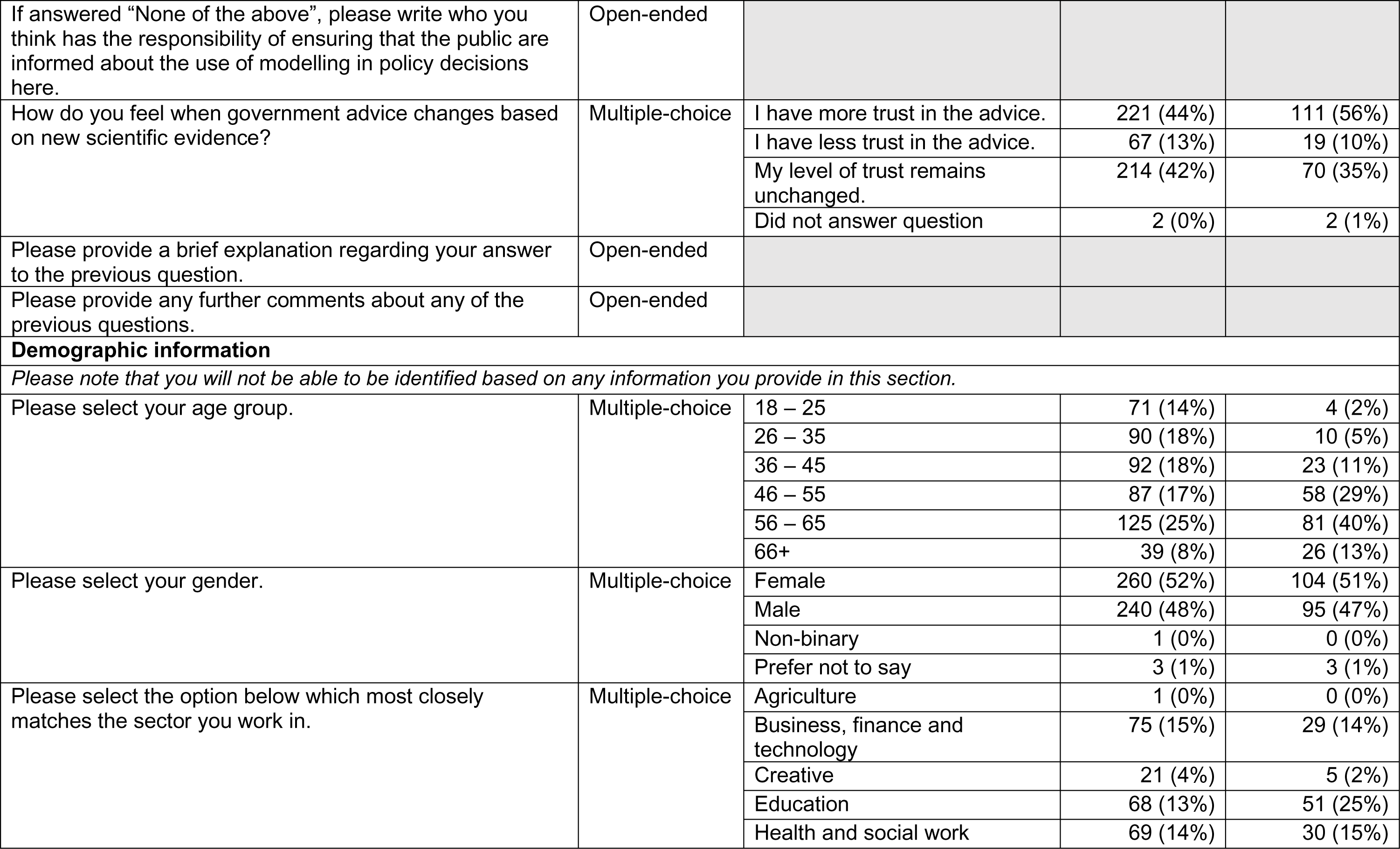
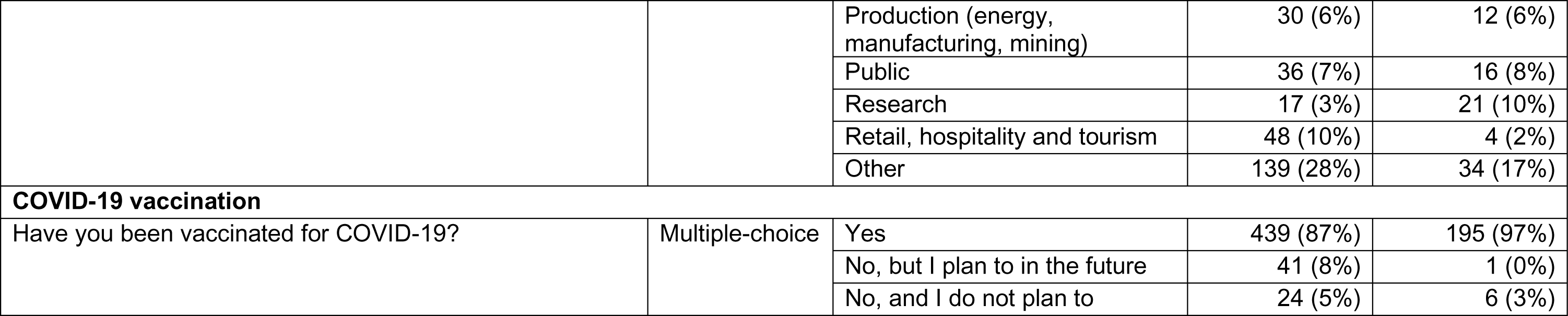
Questions, and multiple-choice answers where relevant, for the survey to the public. The two compulsory questions are highlighted with an asterisk (*). Totals and percentages are based on responses with “complete” demographic data, namely age group, gender, sector and vaccination status within each sample.

To begin, participants were asked to describe a transmission model and state the level of confidence that they have in their description. These were the only two compulsory questions in the survey. Once participants answered this question, a description of transmission models in the context of this survey was given so that answers given in the rest of the survey were relevant to the research questions at hand.

The middle two sections contained most questions which form the basis of the analysis in this paper. Participants were asked a series of questions about their awareness of and opinions on transmission modelling to inform policy before the COVID-19 pandemic. The next section then repeated these questions, also alongside some additional questions, but asked participants to answer in relation to the period during the COVID-19 pandemic.

Finally, participants were asked to provide demographic information, namely age group, gender, the sector that they work in and their COVID-19 vaccination status.

The survey was designed in Microsoft Forms. The questions are presented in full in Table 1.

Participants were gathered via two methods: via an online panel and via social media. Only individuals aged over 18 and residing in the UK were permitted to partake in the survey.

The online panel was surveyed via *Prolific Academic*, an online platform with a large, diverse database of individuals who have consented to participate in research on a multitude of different topics. Our survey was sent to a sample of approximately 500 people signed up to the platform, who were selected to ensure representation across a range of demographic variables, including “Age”, “Sex”, “Country of Birth” and “Employment Status”. As such, this sample is considered a “representative” sample within the context of this paper. Responses were gathered across 7 – 9 July 2021.

Alongside this, we shared our survey on the social media platform *Twitter* by tweeting a link to it, and asked collaborators and accounts associated with the departments to which the authors are affiliated to also do so. An overview of the accounts that tweeted or quote tweeted the survey is presented in Supplementary Table 1. Undoubtedly, a network effect will have influenced who saw the survey in their timeline and who will have selected to participate. Consequently, this sample is considered a non-probability sample. A priori, it was expected that this method of participant recruitment could substantially skew the demographics and answers of the participants within this sample. The survey was first shared on Twitter on 7 July 2021. Although responses were received until 3 September 2021, the vast majority were obtained between 7 – 20 July 2021.

### Statistical analysis

Data were filtered so as to only include observations with all four demographic variables answered (age group; gender; sector; vaccination status). Throughout, percentages of respondents have been rounded to the nearest integer.

#### Classification of open-ended responses

Responses to three open-ended questions, namely “How would you describe a transmission model?”; “Before the COVID-19 pandemic, where do you think those who developed and used transmission models worked?” and “During the COVID-19, where do you think those who developed and used transmission models worked?”, were categorised to aid the interpretation of the results. Categories were not pre-determined and were refined having considered the entirety of the data.

Descriptions of transmission modelling were deemed to be more relevant to the context of the research either in relation to infectious disease epidemiology or to mathematics and statistics more broadly. Each response was considered individually to assess the sentiment of the description. For example, the use of words such as “virus”, “disease”, “infection”, “illness”, “epidemic” or those akin to “COVID-19” (e.g. “coronavirus”, “covid”), or others such as “statistics”, “mathematical”, “data”, “prediction” or “simulation”, were all interpreted as capturing the essence of “disease transmission and control modelling” and thus were considered as “more relevant”, compared to “less relevant”, to the topic at hand.

Answers to the open-ended question about where transmission modellers work (prior to and during the COVID-19 pandemic) were also assessed on an individual basis. If respondents stated multiple places of work, each were categorised individually. While “academia” and references to “university” were classified as “Research (Academia)”, any other reference to “research” or “science” or “laboratories” were classed as “Research (Unspecified affiliation)”. “Public health bodies” encompassed either direct reference to the term “public health” or specific organisations such as the World Health Organization or Public Health England (the United Kingdom Health Security Agency now combines elements of Public Health England and NHS Test and Trace^17^). Responses were classified as “Government” if either that word was used, a specific department was named, such as the “Department of Health” or reference was made to the “civil service”. Any reference to “journalists” or “the press” or “the media” were clustered together under the heading “Media”, but not those stating “Social media” (of which there were extremely few). “Healthcare services” encompassed reference to “health bodies” without direct mention of “public health”. For example, this includes terms such as “hospitals”, “the NHS” and “healthcare providers”. References of “advisory” to government, or “advisers”, including the use of “SAGE”, was classified as “Advisory roles”. All other responses were placed into “Other”. In most instances, answers were classified in more than one category as they were not mutually exclusive.

Responses to the remaining open-ended questions were not explicitly categorised but, as above, were considered on an individual basis.

#### Analysis of answers to multiple-choice questions

Chi-squared tests were used to test for differences in the proportions of respondents selecting specific answers to multiple-choice questions across samples. Within respondents, Wilcoxon Signed Rank tests were used to assess the differences between time periods within respondents. Questions in which there was a discrete scale from one extreme to the other, for example: “Not at all confident” to “Very confident”; or “No trust whatsoever” to “High level of trust”, were enumerated on a scale where 0 represented the middle strength option (e.g., “Moderately confident” or “Moderate level of trust”), with 1 assigned to the highest level (e.g. “Very confident” or “High level of trust”) and −1 assigned to the lowest level (e.g. “Not at all confident” or “No trust whatsoever”) for each time point. Cases with a binary outcome, for example, whether or not a respondent selected a specific means of being aware of transmission modelling (e.g. “Newspaper (online or print)”), were enumerated as 1 if this option was selected and 0 otherwise. Specific details for each question, alongside results tables, are presented in the Supplementary Material.

#### Reliability scores

Answers to the question “On a scale from 1 – 10 with 1 being “extremely unreliable” and 10 being “extremely reliable”, how do you feel about the use of transmission models in informing public health policy”, both “Prior to the COVID-19 pandemic” and “During the COVID-19 pandemic” are referred to as “reliability scores” throughout. This is the only question in which participants were asked for a numeric answer. Mann-Whitney U tests were used to assess differences in scores between samples within time periods, while Wilcoxon Signed Rank tests were used to assess differences in scores between time periods within respondents.

#### Regression analysis

Linear regression was used throughout the study to quantify the relationships between variables. Throughout, identical regression models were fitted separately on data from the online panel and social media samples. Questions with a discrete answer scale were enumerated as described above. Chi-squared tests were used to assess the overall significance of categorical variables in each model. In the main text, results of these tests are reported if they show statistical significance at the 0.05 level. Full results of all linear regression models are presented in the Supplementary Material.

When assessing the relationship between demographic and other variables, due to the large number of employment categories and limited number of respondents selecting that they are unvaccinated, only age group and gender were included within regression models. The youngest age group, 18 – 25 years, and females, the most frequently reported gender, were designated as the reference levels.

## Results

Participants totalled 509 and 207 across the online panel and social media samples, respectively (total participants 716). Of these, 504 (99%) and 202 (98%) had “complete” demographic data and thus were included in the analysis (total participants 706). Table 1 includes the results of the multiple-choice questions by sample. Response rates to multiple-choice question were high in both samples, with at least 98% of respondents with “complete” demographic data answering each question.

Aside from the demographics of the respondents, the results are discussed with respect to four main themes: awareness, reliability, trust in government advice and communication. Extensive further analyses are presented in the Supplementary Material.

### Respondent demographics

Gender had similar distributions across samples (Table 1; Figure 1A), with females holding a slight majority (52% and 51% for the online panel and social media, respectively).

**Figure 1:**
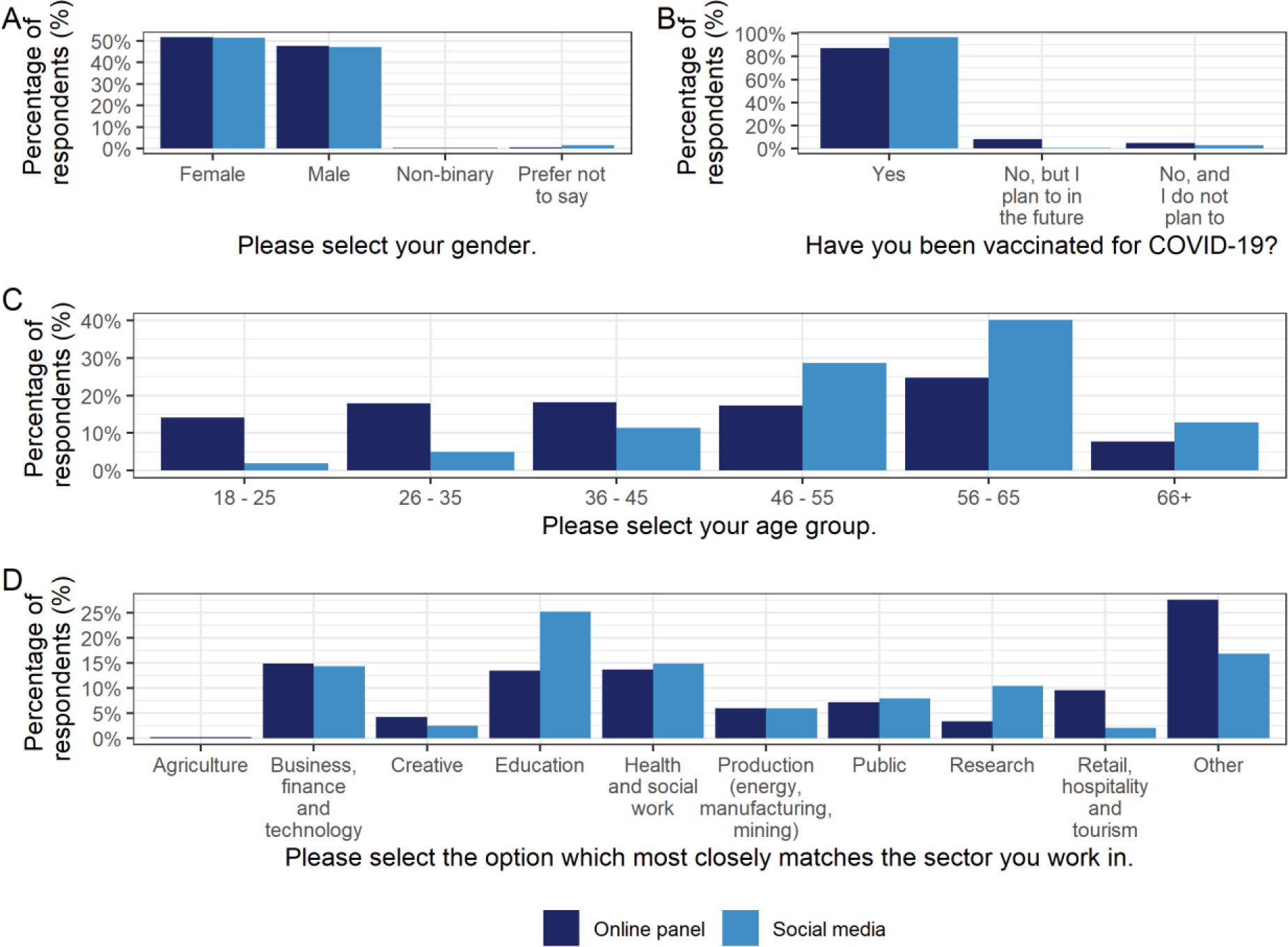
Demographics of survey participants stratified by sample. (A) Gender. (B) COVID-19 vaccination status. (C) Age group. (D) Sector participants work in. Underlying data are presented in Table 1.

There was heterogeneity across the samples in terms of age group and employment sector (Table 1; Figure 1C; Figure 1D). Age groups of the online panel respondents were approximately evenly distributed except for the oldest age group (66+ years) in which there were substantially fewer participants. In contrast, the number in each age group of the social media sample steadily increased up to 56 – 65 years, which had a notably larger percentage of participants than other groups (40%), before also falling in the 66+ category. While half of the employment sector options had fairly consistent percentages of participants working in them across samples, (namely: “Agriculture”, “Business, finance and technology”; “Health and social work”; “Production (energy, manufacturing, mining)”; “Public”), there were fairly large differences elsewhere. For example, social media respondents were significantly more likely than their online panel counterparts to work in “Education” (chi-squared test: 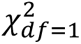 = 13.39; *n* = 706; *p* − *value* < 0.001) or “Research” (chi-squared test: 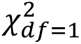 = 12.62; *n* = 706; *p* − *value* < 0.001). This is likely to be attributable to the fact that it was predominantly researchers or research institutions sharing the survey (Supplementary Table 1).

Social media respondents were also significantly more likely to be vaccinated against COVID-19 than online panel respondents (97% and 87%, respectively; Figure 1B; chi-squared test: 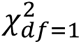 = 13.00; *n* = 706; *p* − *value* < 0.001). However, similar percentage of participants in both samples recording having no intention of being vaccinated (5% and 3%, respectively; chi-squared test: 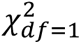 = 0.74, *n* = 706, *p* − *value* = 0.390), with the remaining differences explained by those indicating their intention to be vaccinated in the future.

### Awareness of mathematical transmission modelling

Within samples and time periods, levels of awareness of mathematical transmission modelling (herein referred to as “modelling”) generally were indistinguishable from the levels of awareness of modelling being used in policy specifically (Supplementary Table 2). An exception was online panel respondents in the pre-pandemic period reporting being significantly less aware of modelling being used for policy than modelling in general (Wilcoxon signed rank test: *p* − *value* < 0.001).

The majority of participants across both samples were aware of modelling being used to inform public health policy in the period of the pandemic (Table 1; Figure 2), with these levels being significantly higher than the pre-pandemic period (Online panel: 27% pre-pandemic, 74% during; Wilcoxon signed rank test *p* − *value* < 0.001; Social media: 57% pre-pandemic, 97% during; Wilcoxon signed rank test *p* − *value* < 0.001). However, social media respondents were significantly more likely to be aware of modelling informing policy than online panel respondents within each time period (chi-squared test: prior: 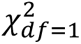 = 56.05; *n* = 706; *p* − *value* < 0.001; during: 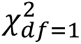 = 44.55; *n* = 706; *p* − *value* < 0.001). Within the social media sample in the pre-pandemic period, males were more likely than females to report awareness of modelling in informing policy (Supplementary Table 3; chi-squared test: *p* − *value* = 0.024).

**Figure 2:**
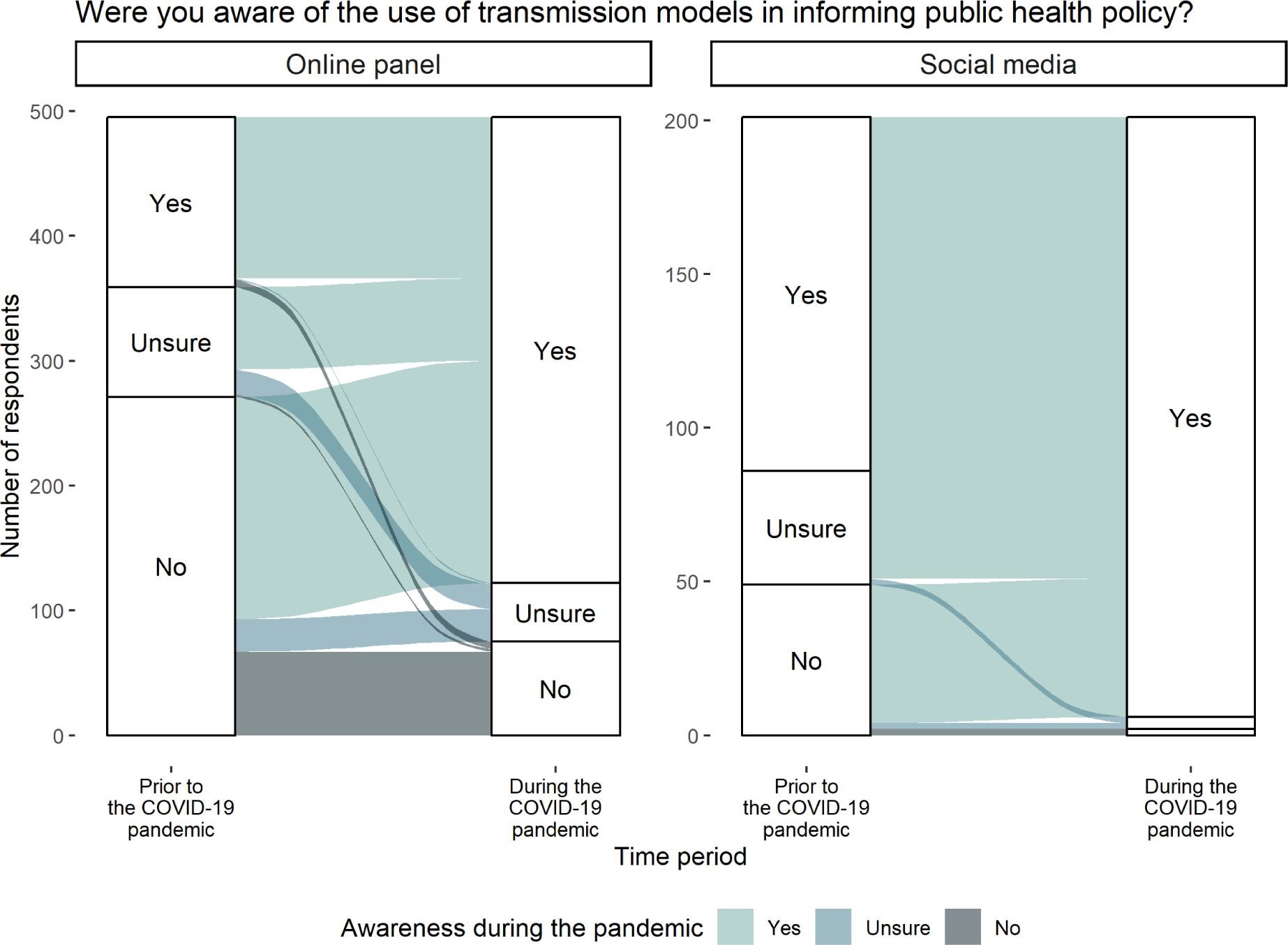
Awareness of modelling informing public health policy. Responses to the question “Were you aware of the use of transmission models in informing public health policy?“ both “Prior to” and “During” the COVID-19 pandemic for the online panel and social media samples. Observations corresponding to a respondent who did not answer the question for at least one time point were removed from the figure to aid visual presentation (n=8 online panel; n=1 social media). Due to the vast majority (97%) of social media respondents reporting being aware of transmission modelling during the COVID-19 pandemic, the options of “Unsure” and “No” are unable to be labelled but appear in the same order as in the three preceding response pillars with “Unsure” above “No”. Underlying data are presented in Table 1.

Online panel respondents were primarily aware of modelling through “News show (TV or online)” or “Newspapers (online or print)” across both time periods (Supplementary Figure 1; Supplementary Table 4). However, social media respondents were primarily aware of modelling through “Academic reports and papers” prior to the COVID-19 pandemic, but “News show (TV or online)”, “Newspapers (online or print)” and “Social media” were the most popular selections during the pandemic. In both time periods, social media respondents were significantly more likely than the online panel to select “Academic reports and papers” (Supplementary Table 5; chi-squared test: prior: *p* − *value* < 0.001; during: *p* − *value* < 0.001), which could be driven in part by the significantly greater percentage of respondents from this sample working in the “Education” and “Research” sectors (Table 1; Supplementary Table 1). Prior to the pandemic, respondents from both samples were equally likely to select “Social media”, but during the pandemic social media respondents were significantly more likely to select this option than their online panel counterparts (Supplementary Table 5; chi-squared test: *p* − *value* < 0.001).

Most participants stated that they had an “About right” level of information of modelling during the pandemic. However, social media respondents were significantly more likely to select this option than their online panel counterparts (51% and 60% of the online panel and social media respondents, respectively; chi-squared test: 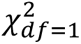 = 4.76; *n* = 706; *p* − *value* = 0.029). Within the social media sample, older age groups were significantly associated with having “too much” information during the pandemic compared to the youngest age group (Supplementary Table 6; chi-squared test: *p* − *value* = 0.018). Some of those selecting the option having an “About right” amount of information also selected that they were unaware of modelling, demonstrating that not all have interest in this topic, in both samples (Supplementary Figure 3; Supplementary Table 7).

### Reliability of mathematical transmission modelling in informing policy

Online panel and social media respondents reported similar mean reliability scores of 6.3 (standard deviation (sd) 1.8) and 6.4 (sd 1.9), respectively, in the pre-pandemic period (Mann-Whitney U test: *p* − *value* = 0.493; Supplementary Figure 4). During the pandemic, mean reliability scores rose significantly to 6.9 (sd 2.0) among online panel respondents (Wilcoxon Signed Rank test: *p* − *value* < 0.001), but rose less among the social media respondents (mean 6.7 (sd 2.3); Wilcoxon Signed Rank test: *p* − *value* = 0.204). There was no evidence to suggest significant differences between mean reliability scores across the two samples during the pandemic (Mann-Whitney U test: *p* − *value* = 0.613).

A majority of respondents changed their reliability score across time periods, with social media respondents being significantly more like to do so (Figure 3; Supplementary Table 11; online panel 55%; social media 65%; chi-squared test: 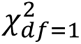 = 4.96; *n* = 706; *p* − *value* = 0.026). Respondents were similarly more likely to perceive greater reliability during the pandemic compared to before, regardless of sample (40% and 39% for online panel and social media, respectively; 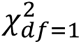 = 0.04 *n* = 706; *p* − *value* = 0.840). However, social media respondents were around 70% more likely to have a decreased feeling of reliability compared to their online panel counterparts (26% and 15% for social media and online panel, respectively; 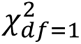 = 10.34; *n* = 706; *p* − *value* = 0.001).

**Figure 3:**
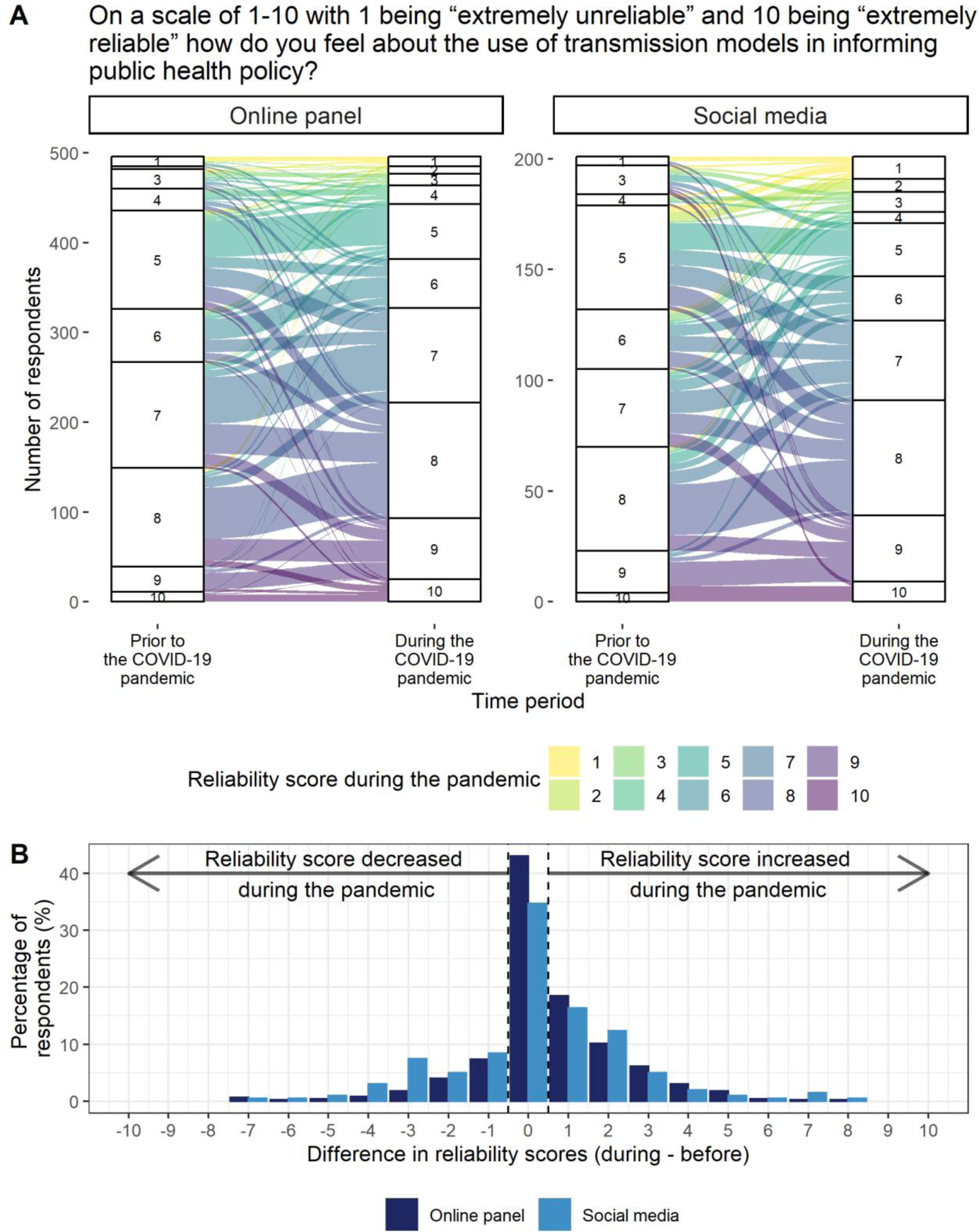
Reliability of using transmission modelling to inform public health policy. (A) Responses to the question “On a scale of 1 – 10 with 1 being “extremely unreliable” and 10 being “extremely reliable” how do you feel about the use of transmission modelling in informing public health policy?” both “Prior to” and “During” the COVID-19 pandemic, for both the online panel and social media samples. Observations corresponding to a respondent which did not answer the question for at least one time point were removed from the figure to aid visual presentation (n=8 online panel; n=1 social media). Due to small numbers of respondents from the online panel selecting a score of “2”, this is unable to be labelled, but appears in numeric order. Underlying data are presented in Table 1. (B) Distribution of the within-respondent differences in reliability scores from during the pandemic compared to prior for both the online panel and social media samples. Difference in scores is defined as the reliability score during the pandemic minus reliability score prior to the pandemic.

During the COVID-19 pandemic, both older age groups and being male were significantly associated with lower reliability scores compared to younger age groups and females, respectively within the social media sample only (Supplementary Table 8; chi-squared tests: age: *p* − *value* = 0.031; gender: *p* − *value* = 0.001). Awareness was associated with a similar reliability score within this sample during the pandemic (*p* − *value* = 0.817). In contrast, within the online panel at both time points and social media in the pre-pandemic period, increased awareness of the use of modelling in informing policy was associated with a significant increase in feeling of reliability rather than the demographic variables (Supplementary Table 9; online panel and pre-pandemic: *p* − *value* < 0.001; online panel and during: *p* − *value* < 0.001; social media and pre-pandemic: *p* − *value* = 0.024). However, only 4% of social media respondents reported not being aware during the pandemic, which is an insufficient sample size from which to draw robust conclusions.

### Trust in government advice

Most respondents reported having “moderate” trust in the government regarding public health issues, regardless of sample or time period (Table 1; Figure 4A). Particularly in the pre-pandemic period, many respondents stated their belief that the government had the best interest of the public at heart, with one stating “I didn’t have [a] reason not to” (online panel). Within both samples, respondents noted that they had a high level of trust in scientific evidence rather than the government: “I trusted government advice because this is provided by scientific advisors” (online panel) and “I trusted the scientist[s] not the politicians” (social media), acknowledging that decisions taken by government can be influenced by more than just science: “It can be difficult to disentangle the science from politics” (social media). The perception of the government not trusting scientific advice was frequently mentioned as a reason for a lower level of trust, for example: “They didn’t always listen to scientists, which deteriorated my trust” (online panel). These views were reflected in the reliability scores with a higher level of trust in government advice being associated with higher median reliability scores for both samples. However, there was substantially greater variance in reliability scores among those with lower trust levels. (Supplementary Figure 9; Supplementary Table 13).

**Figure 4:**
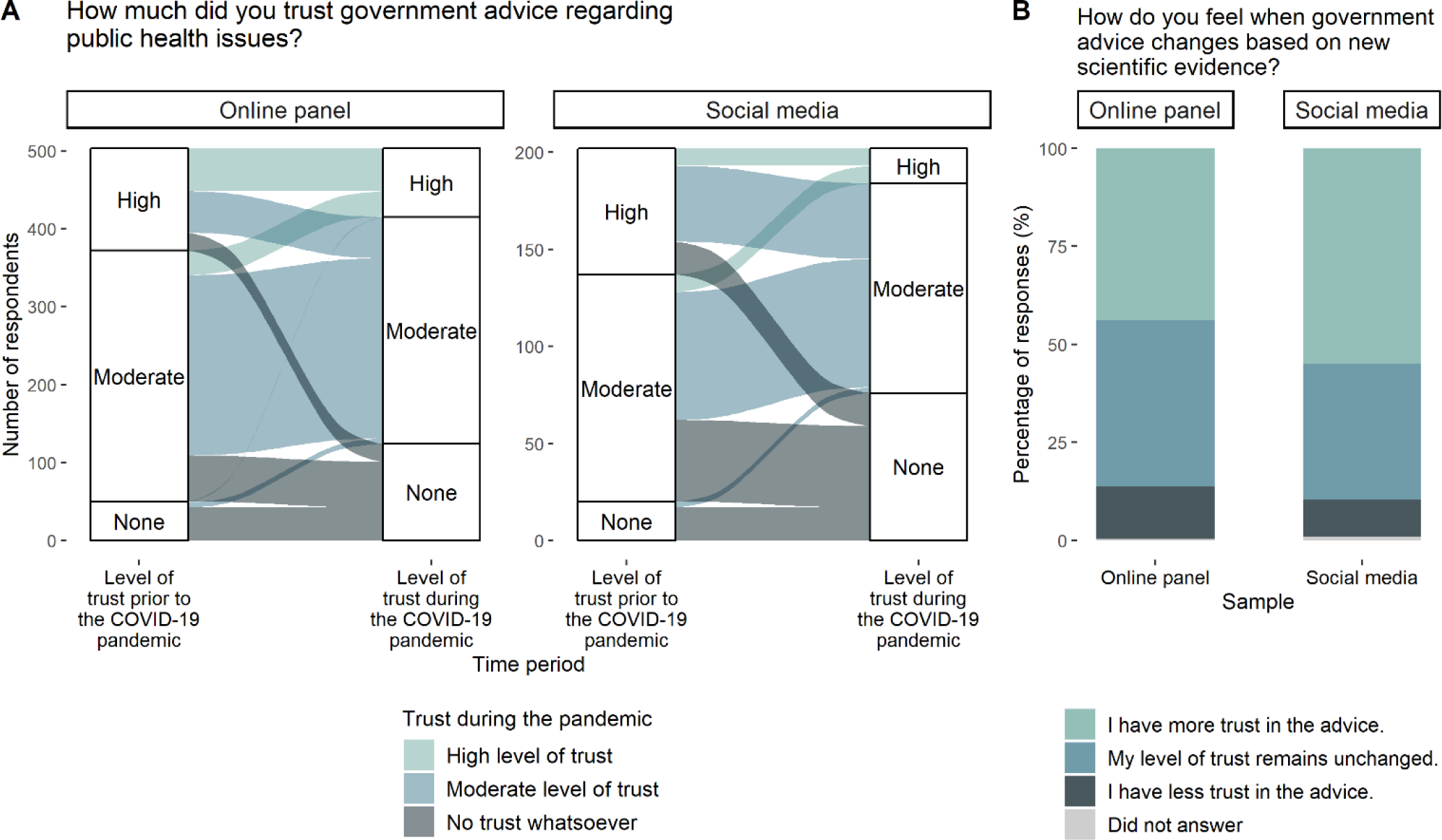
Trust in government scientific advice. (A) The percentage of observations from both the online panel and social media samples according to time period and answer to the question “How much did you trust government advice regarding public health issues?” both “Prior to” and “During” the COVID-19 pandemic. Observations corresponding to respondents who did not answer the question for at least one time point were removed from the figure to aid visual presentation (n=1 respondent from the online panel). Underlying data are presented in Table 1. (B) Answers to the question “How do you feel when government advice changes based on new scientific evidence?” for both the online panel and social media samples. Underlying data are presented in Table 1.

Most social media respondents reported a different level of trust in government public health advice in the period of the pandemic compared to the pre-pandemic period (54%), significantly more than the minority of online panel respondents also doing so (35%) (chi-squared test: 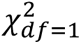 = 22.51; *n* = 706; *p* − *value* < 0.001) (Supplementary Table 14). In particular, the percentage of respondents selecting having “No trust whatsoever” rose significantly during the pandemic from 10% (both samples) to 25% and 38% for online panel and social media respondents, respectively (Wilcoxon signed rank test: Online panel: *p* − *value* < 0.001; Social media: *p* − *value* < 0.001). 26% of social media respondents and 18% of online panel respondents went from a “High level of trust” to “No trust whatsoever” during the pandemic (chi-squared test: 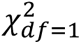 = 1.48; *n* = 706; *p* − *value* = 0.223). Reasons given for this included: “Felt that they [the government] were more driven by political and economic priorities rather than public health.” (online panel) and “Not convinced they followed the science on every occasion as they stated they were doing.” (social media). Separately, another respondent noted: “My trust has reduced but not yet to the position of no trust” (online panel). Together, this demonstrates an erosion of trust among respondents, but substantially more so among the social media sample.

One respondent who went from “High” to “No” trust gave the following reason for their opinion: “Government kept changing the advice given during the pandemic.” (online panel). However, when asked directly about trust in relation to the government changing their advice based on scientific evidence, a minority of respondents from both samples stated consequently having “Less trust” (Table 1; Figure 4B; 13% and 10% for online panel and social media, respectively; chi-squared test: 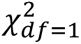 = 1.69, *n* = 706; *p* − *value* = 0.194). Among those stating they find changing advice “Less trustworthy”, respondents noted: “It makes me feel as though the government make up things to give the public false hope until there is scientific research available to support” (online panel) and “Gets too confusing [with] mixed messages” (social media). Others among the online panel sample suggested that it was the rapid pace at which advice changed which influenced their level of trust: “the advice changed so frequently it was hard to know what to believe was the right thing to do”.

Social media respondents (56%) were “More trusting” of changing advice based on new evidence than their online panel counterparts (44%) (chi-squared test: 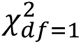 = 6.70; *n* = 706; *p* − *value* = 0.010), although this was still the most common response among both samples. Respondents noted: “It shows a willingness to address a changing situation” (online panel) and “During a pandemic there will be need to amend advice as more data becomes available and more [is] known about the virus” (social media). Across both samples, it was clear that respondents placed a high value on understanding the source of the new information (“More trust as long as sources are quoted” (online panel)) and the method in which it was explained to the public (“I understand that evidence as it gathered can lead to new understanding and therefore improved modelling. However, it can be dependent on how well it is explained and communicated.” (social media)).

Being male was significantly associated with a lower level of trust in changing advice compared to being female within the social media sample (Supplementary Table 19; chi-squared test: 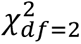 = 2.75; *n* = 202; *p* − *value* = 0.045). Within the online panel only, awareness of the use of modelling in informing policy during the pandemic was associated with greater trust in changing scientific advice (Supplementary Table 20; *p* − *value* = 0.049), as were higher levels of trust in government scientific advice during the pandemic (Supplementary Table 21; *p* − *value* = 0.024).

### Responsibility of communicating mathematical transmission modelling to the public

Almost all respondents believed that governments have the responsibility of ensuring that the public are informed about the use of modelling in informing policy decisions (92% of online panel respondents; 91% of social media respondents; Table 1; Supplementary Figure 13; chi-squared test: 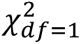 = 0.10; *n* = 706; *p* − *value* = 0.756). Social media respondents were significantly more likely to select those who “develop” or “use” transmission models, “The media”, and “Myself, as a member of the public” as having the responsibility than online panel respondents (Supplementary Table 23). Except for “Myself, as a member of the public”, the three aforementioned responses were equally popular within each sample (Supplementary Table 23).

## Discussion

This study was undertaken to complement the findings of our previously published work on the relationship between modelling and policy^2^ by providing insights into the UK public’s awareness and opinion of transmission modelling in informing policy. As such, novel data were gathered under two different sampling mechanisms, an online panel (“representative” sample) and social media (“non-probability” sample) and across two time periods, “Prior to” and “During” the COVID-19 pandemic.

Almost all respondents were increasingly aware of the use of modelling in informing policy during the pandemic, with significantly higher levels of awareness among social media respondents than online panel respondents. Awareness generally stemmed from the news media and social media during the pandemic. The mean reliability score increased during the pandemic compared to the pre-pandemic period under both samples, with awareness being positively associated with reliability within both samples and time points, except for social media during the pandemic. Trust in government public health advice remained high across samples and time periods overall but was lower in the period of the pandemic compared to the pre-pandemic period. The decay in trust was notably greater among social media respondents. Many respondents from both samples explicitly made the distinction that their trust was reserved for “scientists” and not “politicians”. Almost all respondents, regardless of sample, believed governments have responsibility for the communication of modelling to the public.

Our results are not, nor claim to be, a definitive representation of all public opinion on this topic. Instead, we have focussed primarily on the differences in the responses between our two samples. Specifically, our results validate our a priori expectation of a skewed response among social media respondents thus providing an important reminder of the potentially biased conclusions that could be drawn from non-representative samples.^18^ Throughout, it is important to recall that social media respondents were likely to have accessed the survey directly from a Twitter account they follow related to public health or mathematical modelling, thus demonstrating an active interest in and/or familiarity with the subject matter (Supplementary Table 1). In contrast, online panel respondents saw this survey without any context. Furthermore, social media respondents were significantly more likely to work in “Research” or “Education” than online panel respondents, which may account for a substantial amount of the differences observed in the responses received, given that modelling for public health policy was primarily undertaken by academics.^5^

While the general sentiment of the responses was consistent across samples, often social media respondents were more likely to have stronger or polarised views. Examples of this include trust in the government decreasing within both samples but at a greater level for social media respondents compared to the online panel, and almost all social media respondents stating awareness of the use of modelling in informing policy compared to three quarters of online panel respondents. Except for one question, age and gender were only found to be significantly associated with responses among the social media sample. Nonetheless, caution is still needed when interpreting the results from the online panel as the suitability of this sample as a representation of the UK population is unclear. However, 87% of online panel respondents (aged over 18 years old) stated having had at least one dose of a vaccine for SARS-CoV-2 compared to 76.9% of the population over 12 years old in the UK on 9 July 2021 (when online panel survey responses were gathered).^19^ As vaccination of those aged under 18 years was not recommended in the UK until 15 July 2021^20^, it is reasonable to consider 76.9% a lower bound of vaccination coverage of the UK adult population thus suggesting reasonable representativeness of the online panel with regards to this variable.

Given the role that modelling has played in the pandemic response in the UK,^2, 3, 5, 6^ and the corresponding coverage in the media,^21–24^ it is unsurprising that respondents reported an increased awareness of the use of modelling as a tool to inform public health policy. However, it is perhaps unexpected that this translated to most participants in both samples. The popularity of the news media as a means of awareness underlines the important role that journalists play as science communicators, and emphasises the value of organisations such as the Science Media Centre.^25^ A number of the experts interviewed as part of our aforementioned study praised the commitment that many journalists showed to clear and accurate communication of modelling studies.^2^ The rise in popularity of social media as a means for being aware of modelling is unsurprising among social media respondents, but the significant four-fold increase among online panel users in the period of the pandemic was particularly notable. Akin to the case of the news media, this finding further demonstrates the role that social media can play in science communication, particularly in the COVID-19 pandemic, and, consequently, the importance of ensuring that studies are properly represented to users of this platform by those sharing them.

One of the major findings of the authors’ initial science-policy paper was the importance of distinguishing between scientists and decision-makers, with only the latter being directly accountable to the public.^2^ Many respondents, from both samples, noted this distinction explicitly when commenting on the question regarding trust in government public health policy, and within both samples during the pandemic respondents were significantly more likely to state that modellers work within “Academia” rather than “Government” (see Supplementary Material). While this suggests a reasonable understanding of the difference between these two roles, further and continued emphasis of the difference in roles may be warranted ahead of future emergencies, public health-related or otherwise, as “Government” became a significantly more popular response during the pandemic compared to the pre-pandemic period among the online panel. Nearly all respondents from both samples indicated that the government have the responsibility of ensuring that the public are informed of scientific evidence underpinning their decisions. This remains compatible with another key finding of our initial study that science must be communicated by scientists,^2^ for example by having scientists present at Government-organised press conferences.

There will often be tensions between scientific advice and decision-making. This was particularly evident during the COVID-19 pandemic when our understanding of SARS-CoV-2 was rapidly evolving as new data became available, with public health guidance changing almost equally rapidly to reflect new findings. This was highlighted by science communication experts in our previous study as a key challenge of communicating with the public and was encapsulated well by a social media respondent: “Politicians don’t like u-turns while scientists are keen to constantly evolve thinking and theories. These outlooks are not particular[ly] compatible when it comes to communicating subtle changes in public health messaging.”. However, most respondents from both samples noted that changing advice either did not change or increased their trust in said advice, although this was significantly more common among social media respondents. Our results indicate a key factor of trust is transparency or evidence, also highlighted in our initial study,^2^ rather than rapidly changing advice.

The fact that respondents reported “No awareness” of modelling in informing policy and that for some (online panel *n* = 10; social media *n* = 1) this was an “About right” level of knowledge provides an important reminder that this topic is, understandably, not of interest to all. Furthermore, while awareness was associated with increased feelings of reliability of modelling, the fact that some “Aware” respondents selected a reliability score in the lower half of the scale during the pandemic highlights the complexities of this relationship. Nonetheless, some respondents seemed to show a genuine interest and appreciation of modelling: “I have found a new respect for those who do modelling, to be honest I find it fascinating” (online panel).

It is important to note the limitations of this study. All results rely on the opinions and experiences of our participants. Although we distributed the survey via two platforms, as discussed we are unable to ascertain the representativeness of the online panel in terms of the UK population. Furthermore, although respondents were asked questions in relation to the period “Prior to” and “During” the COVID-19 pandemic separately, sampling only took place during the latter period. Therefore, responses for the pre-pandemic period may have been influenced by recall bias and should be interpreted with caution. Due to the nature of the sampling platforms, all online panel responses were obtained in a much shorter period (days) than social media responses (weeks). Around this time, all public health restrictions in England were lifted after a one-month delay^26^, which could have influenced participants’ responses. Finally, although respondents were given multiple opportunities to add open-ended comments, multiple-choice answers may lose some of the nuance of participants’ opinions.

As noted by one respondent: “I think we have all learned something about science and how it impacts on our daily lives throughout the pandemic”. Ultimately, the public are key stakeholders of public health policy, and if mathematical transmission models are used to inform important decisions then there is a responsibility to assess people’s understanding of and feelings towards them. We hope that the findings of this study will be useful in informing science communication strategies in future emergencies.

## Ethics

This study has approval from the Medical Sciences Interdivisional Research Ethics Committee at the University of Oxford with Ethics Approval Reference R76166/RE001.

## Data and code availability

In line with the ethics approval obtained, only RM and CAD have access to the raw research data. However, anonymised summary data are provided in Table 1. Code for R analyses are provided at https://github.com/ruthmccabe/public-survey-modelling.

## Author contributions

RM and CAD designed the study. RM conducted the analysis and wrote the first draft of the manuscript. CAD edited the manuscript.

## Acknowledgements

This work was supported by the NIHR Health Protection Research Unit (HPRU) in Emerging and Zoonotic Infections, a partnership between PHE, University of Oxford, University of Liverpool and Liverpool School of Tropical Medicine [grant number NIHR200907 supporting RM and CAD]; and the MRC Centre for Global Infectious Disease Analysis [grant number MR/R015600/1 supporting CAD], which is jointly funded by the UK Medical Research Council (MRC) and the UK Foreign, Commonwealth and Development Office (FCDO), under the MRC/FCDO Concordat agreement and is also part of the EDCTP2 programme supported by the European Union (EU).

Disclaimer: “The views expressed are those of the authors and not necessarily those of the United Kingdom (UK) Department of Health and Social Care, EU, FCDO, MRC, National Health Service, NIHR, or PHE. The funding bodies had no role in the design of the study, analysis and interpretation of data and in writing the manuscript.”

The authors would like to thank a number of individuals who supported this analysis. First, to Professor Sally Sheard, Dr Hayley Mableson and Dr Michael Humann for their helpful comments when drafting the survey. Second, to all those who shared our survey via social media. Third, to Cathal Mills at the University of Oxford for his assistance with data visualisation. Last, but by no means least, the authors are incredibly grateful to all those who took the time to complete the study.

