## Supplementary Material for "Public awareness of and opinions on the use of mathematical transmission modelling to inform public health policy in the United Kingdom"

Ruth McCabe[1,2,3,*] and Christl A. Donnelly[1,2,3,4]

[1] Department of Statistics, University of Oxford

[2] NIHR Health Protection Research Unit in Emerging and Zoonotic Infections

[3] Pandemic Sciences Institute, University of Oxford

[4] MRC Centre for Global Infectious Disease Analysis, Imperial College London

### Participant Information Sheet

Department of Statistics
24-29 St Giles'
Oxford
OX1 3LB

Professor Christl Donnelly
University Direct Line: 01865 ******
University

UNIVERSITY OF OXFORD

**Awareness and opinions on the use of transmission modelling in informing policy**

CUREC Approval Reference: R76166/RE001

#### *General Information*

We recently wrote an article for a special edition of the *Journal of the Royal Society Interface Focus* themed on COVID-19 in society. We looked at this through the lens of mathematical modelling. We surveyed attendees of the Scientific Advisory Group for Emergencies (SAGE) or the Scientific Pandemic Influenza Group on Modelling (SPI-M), meetings to elicit their experiences of the development and use of transmission modelling in policy throughout the COVID-19 pandemic. We now want to build upon this work by surveying the public on their awareness and opinions of the use of modelling in informing policy throughout the pandemic. Please note that participants are not required to have any prior knowledge of transmission modelling to participate.

We appreciate your interest in participating in this online survey. You have been invited to participate as you are an adult over 18 years of age and residing within the United Kingdom of Great Britain and Northern Ireland.

You may ask any questions by contacting the researcher (details below).

The Principal Researcher is Ruth McCabe, who is attached to the Department of Statistics at the University of Oxford. This project is being completed under the supervision of Professor Christl Donnelly.

You are invited to participate in a survey consisting of 13 multiple choice questions and 8 open-ended questions. At the end of the survey, basic demographic information covering age, gender and occupation are asked for. Participants cannot be identified from this information. This should take about 10 minutes.

The data gathered will be used to write a short commentary in follow up to our original article and will also be incorporated into the corresponding doctoral thesis in statistics. Answers to all questions are anonymous. Only the Principal Researcher and Supervisor will have access to the raw, anonymised data.

#### *Do I have to take part?*

No. Please note that participation is voluntary. If you do decide to take part, you may withdraw at any point for any reason before submitting your answers by closing the browser. However, once submitted your data cannot be removed due to their being no identifiable information collected.

#### *How will my data be used?*

We will not collect any information that could be used to identify you.

Anonymous research data (including consent records) will be stored for three years after publication or public release.

Your IP address will not be stored. We will take all reasonable measures to ensure that data remain confidential.

The responses you provide will be used in an academic publication.

#### *Who will have access to my data?*

The researchers named above will have access to the raw data that you provide, which is anonymised and from which you cannot be identified. The University will process the data you provide for the purpose of the research outlined above. Research is a task that we perform in the public interest. Further information about your rights with respect to your personal data is available from https://compliance.admin.ox.ac.uk/individual-rights.

The data you provide may be shared with third parties (researchers at partner universities) for analysis and publication purposes.

This survey will be written up for a doctoral thesis in statistics.

#### *Who has reviewed this study?*

This project has been reviewed by, and received ethics clearance through, the Medical Sciences Interdivisional Research Ethics Committee, a subcommittee of the University of Oxford Central University Research Ethics Committee R76166/RE001.

#### *Who do I contact if I have a concern or I wish to complain?*

If you have a concern about any aspect of this study, please write to Ruth McCabe at or her supervisor Professor Christl Donnelly at, and we will do our best to answer your query. We will acknowledge your concern within 10 working days and give you an indication of how it will be dealt with. If you remain unhappy or wish to make a formal complaint, please contact the Chair of the Medical Sciences Interdivisional Research Ethics Committee at the University of Oxford who will seek to resolve the matter as soon as possible:

; Address: Research Services, University of Oxford, Wellington Square, Oxford OX1 2JD

The following two statements form the first two questions of the survey, which you must complete in order to proceed to the rest of the questions.

**Please note that you may only participate in this survey if you are 18 years of age or over and residing in the United Kingdom of Great Britain and Northern Ireland (UK).**

☐ I certify that I am 18 years of age or over and reside in the UK.

**If you have read the information above and agree to participate with the understanding that the data you submit will be processed accordingly, please check the relevant box below to get started.**

☐ Yes, I agree to take part

### Methods

Supplementary Table 1 presents the details of the accounts who shared the link to the survey, either by tweeting it directly or quote tweeting the link, as of 18 July 2023.

***Supplementary Table 1:*** *Details of accounts who directly tweeted or quote tweeted (QT) the link to the survey.*

| **Type of tweet** | **Account name** | **Twitter handle** | **Date tweeted** | **Link** | **Number of quote tweets** | **Number of retweets** | **Number of likes** | **Number of replies** |
|---|---|---|---|---|---|---|---|---|
| Direct | HPRU EZI | @HPRUezi | 7 July 2021 | [1] | 3 | 6 | 5 | 4 |
| Direct | Oxford Statistics | @Oxford Stats | July 2021 | [2] | Although we have a link to this Tweet, it is unfortunately no longer available and so specific information on reach is unavailable. | | | |
| QT | Liam McCabe | @liamrun | 7 July 2021 | [3] | 0 | 0 | 1 | 0 |
| QT | Ruth McCabe | @ruth_ mccabe | 16 July 2021 | [4] | 3 | 6 | 6 | 1 |
| QT | Jameel Institute | @Imperial _Jameel | 16 July 2021 | [5] | 0 | 1 | 6 | 0 |
| QT | Dr David Telford | @Davie_T | 16 July 2021 | [6] | 0 | 1 | 4 | 0 |
| QT | Marie | @marie clarkg12 | 16 July 2021 | [7] | 0 | 1 | 3 | 2 |
| QT | Katharina Hauck | @kdhauck | 16 July 2021 | [8] | 0 | 0 | 6 | 0 |
| QT | Oonagh Gil | @Oonagh Gil | 17 July 2021 | [9] | 0 | 1 | 3 | 0 |
| QT | MRC Centre for Global Infectious Disease Analysis | @MRC_ Outbreak | 20 July 2021 | [10] | 0 | 2 | 3 | 0 |
| QT | David Spiegelhalter | @d_ spiegel | 2 August 2021 | [11] | 0 | 9 | 24 | 2 |

[1] https://twitter.com/HPRUezi/status/1412793471895748608
[2] https://twitter.com/OxfordStats/status/1413446810111135753
[3] https://twitter.com/liamrun/status/1412819957696638976
[4] https://twitter.com/ruth_mccabe/status/1415943038900264962
[5] https://twitter.com/Imperial_Jameel/status/1415984380045152256
[6] https://twitter.com/Davie_T/status/1415958167687733251
[7] https://twitter.com/marieclarkg12/status/1415988394447020033
[8] https://twitter.com/kdhauck/status/1416000017597358080
[9] https://twitter.com/OonaghGil/status/1416347577730146306
[10] https://twitter.com/MRC_Outbreak/status/1417485287932956673
[11] https://twitter.com/d_spiegel/status/1422190746787106817

### Results: Figures, tables and additional analyses

#### *Awareness of mathematical transmission modelling*

#### Were you aware of the use of transmission models in informing public health policy?

Supplementary Table 2 presents the results of Wilcoxon signed rank tests for differences between awareness of modelling generally and awareness specifically in relation to use in policy, for both samples and time periods. Responses in which a respondent stating having no awareness or being unsure were enumerated as 0, with responses stating awareness enumerated as 1.

Supplementary Table 3 presents the results of the linear model regressing responses to the question "Were you aware of the use of transmission models in informing public health policy?" both "Prior to" and "During" the COVID-19 pandemic on age group and gender for each sample. The three responses provided as multiple-choice options, "No", "Unsure" and "Yes", were enumerated as -1, 0, 1, respectively.

***Supplementary Table 2:*** *Differences between awareness of transmission modelling generally and specifically in relation to use in policy for both time periods and samples according to Wilcoxon signed rank tests.* $p - values < 0.05$ *are considered statistically significant.*

| | | **Number of respondents (n (%))** | | **Wilcoxon signed rank test statistic** | $n$ | $p - value$ |
|---|---|---|---|---|---|---|
| | | **Not aware generally** | **Not aware or unsure regarding policy** | | | |
| **Prior to the COVID-19 pandemic** | **Online panel** | 314 (62%) | 365 (72%) | 3927 | 504 | <0.001 |
| | **Social media** | 74 (37%) | 86 (42%) | 581 | 202 | 0.065 |
| **During the COVID-19 pandemic** | **Online panel** | 114 (23%) | 122 (24%) | 1326 | 504 | 0.180 |
| | **Social media** | 6 (3%) | 6 (3%) | 18 | 202 | 1.000 |

***Supplementary Table 3:*** *Coefficient estimates, standard errors and p-values for the linear model regressing responses to the question "Were you aware of the use of transmission models in informing public health policy?" both "Prior to" and "During" the COVID-19 pandemic on age group and gender for each sample. Chi-squared tests for the overall significance of age and gender are also presented. The three responses provided as multiple-choice options, "No", "Unsure" and "Yes", were enumerated as -1, 0, 1, respectively. $p - values < 0.05$ are considered statistically significant.*

| Sample | Response | Variable | Interpretation | Estimate | Standard Error | p-value | Chi-squared test | |
|---|---|---|---|---|---|---|---|---|
| | | | | | | | $\chi^2$ | p-value |
| Online panel | Prior to the COVID-19 pandemic, were you aware of the use of transmission models in informing public health policy? | Intercept | Baseline* | -0.42 | 0.11 | <0.001 | | |
| | | Age (years) | 26 – 35 | 0.17 | 0.14 | 0.227 | 3.63 (df=5) | 0.432 |
| | | | 36 – 45 | 0.05 | 0.14 | 0.725 | | |
| | | | 46 – 55 | 0.20 | 0.14 | 0.140 | | |
| | | | 56 – 65 | 0.06 | 0.13 | 0.650 | | |
| | | | 66+ | -0.13 | 0.17 | 0.465 | | |
| | | Gender | Male | 0.14 | 0.08 | 0.072 | 3.56 (df=3) | 0.189 |
| | | | Non-binary | -0.75 | 0.87 | 0.387 | | |
| | | | Prefer not to say | -0.34 | 0.50 | 0.495 | | |
| Social media | Prior to the COVID-19 pandemic, were you aware of the use of transmission models in informing public health policy? | Intercept | Baseline* | 0.29 | 0.43 | 0.501 | | |
| | | Age (years) | 26 – 35 | 0.14 | 0.49 | 0.774 | 2.01 (df=5) | 0.718 |
| | | | 36 – 45 | 0.07 | 0.45 | 0.882 | | |
| | | | 46 – 55 | -0.19 | 0.43 | 0.669 | | |
| | | | 56 – 65 | -0.13 | 0.43 | 0.771 | | |
| | | | 66+ | -0.10 | 0.45 | 0.823 | | |
| | | Gender | Male | 0.28 | 0.12 | 0.019 | 5.30 (df=2) | 0.024 |
| | | | Prefer not to say | 0.84 | 0.50 | 0.093 | | |
| Online panel | During the COVID-19 pandemic, were you aware of the use of transmission models in informing public health policy? | Intercept | Baseline* | 0.51 | 0.01 | <0.001 | | |
| | | Age (years) | 26 – 35 | 0.01 | 0.12 | 0.927 | 2.11 (df=5) | 0.569 |
| | | | 36 – 45 | 0.03 | 0.12 | 0.792 | | |
| | | | 46 – 55 | 0.18 | 0.12 | 0.140 | | |
| | | | 56 – 65 | 0.13 | 0.11 | 0.223 | | |
| | | | 66+ | 0.11 | 0.15 | 0.458 | | |
| | | Gender | Male | 0.03 | 0.07 | 0.694 | 0.83 (df=3) | 0.679 |
| | | | Non-binary | 0.48 | 0.74 | 0.517 | | |
| | | | Prefer not to say | 0.43 | 0.43 | 0.313 | | |
| Social media | During the COVID-19 pandemic, were you aware of the use of transmission models in informing public health policy? | Intercept | Baseline* | 0.94 | 0.12 | <0.001 | | |
| | | Age (years) | 26 – 35 | -0.19 | 0.14 | 0.185 | 0.38 (df=5) | 0.247 |
| | | | 36 – 45 | -0.06 | 0.13 | 0.623 | | |
| | | | 46 – 55 | -0.01 | 0.12 | 0.940 | | |
| | | | 56 – 65 | 0.01 | 0.12 | 0.962 | | |
| | | | 66+ | -0.01 | 0.13 | 0.921 | | |
| | | Gender | Male | 0.08 | 0.03 | 0.023 | 0.30 (df=2) | 0.074 |
| | | | Prefer not to say | 0.05 | 0.14 | 0.708 | | |

*Baseline comprised of "Female" and "18 – 25" years.

#### How were you aware of mathematical transmission modelling?

Supplementary Figure 1 presents responses to the question "How were you aware of transmission modelling?" for both samples and for both time points.

Supplementary Table 4 presents the results of Wilcoxon signed rank tests to assess differences in the number of respondents selecting responses to the aforementioned questions across the two time periods. For each multiple-choice answer, data were enumerated to a binary scale with 1 indicating that this answer was selected and 0 indicating that it was not selected.

Supplementary Table 5 present the results of chi-squared tests for differences between the proportions of respondents selecting responses to the aforementioned question across the two samples.

Although not among the most common responses, the percentage of respondents reporting "Internet search" as their source of information on modelling almost doubled and more than tripled in the period during the pandemic compared to the period before, for the online panel (Wilcoxon signed rank test $p-value < 0.001$) and social media (Wilcoxon signed rank test $p-value < 0.001$) respectively, indicating an increased appetite for information on mathematical modelling among both samples. This was verified by individuals commenting that their trust is often highly dependent on transparency of evidence, which was also highlighted in our initial study.

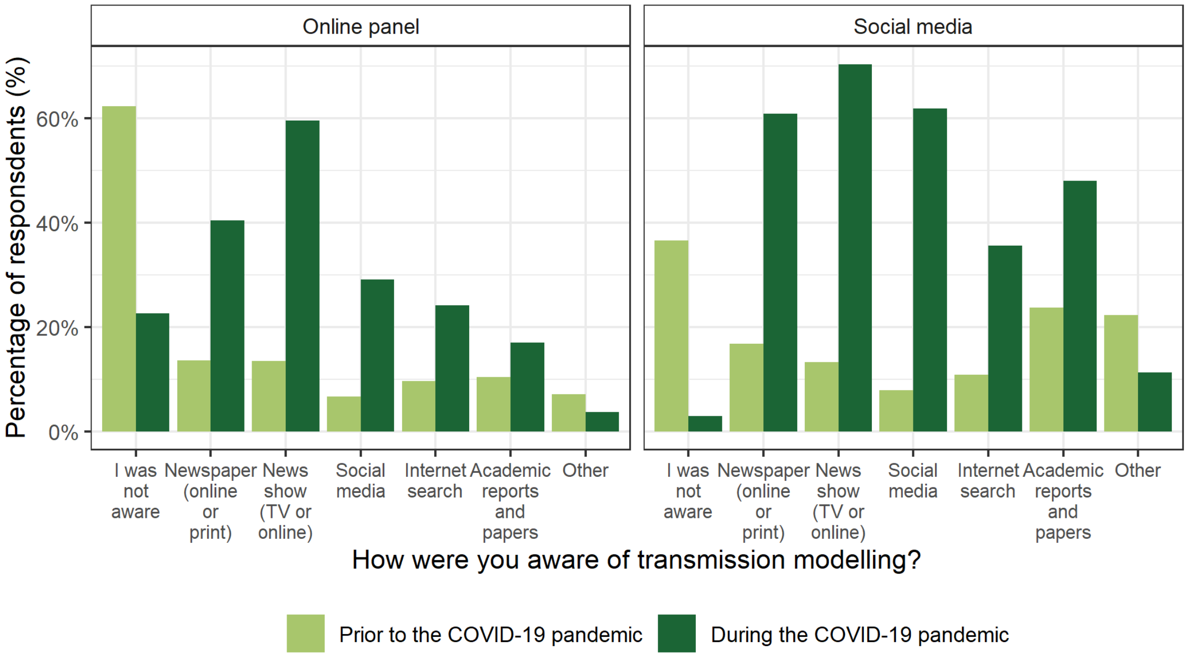

***Supplementary Figure 1:*** *Responses to the question "How were you aware of transmission modelling?" either "Prior to" or "During" the COVID-19 pandemic for both the online panel and social media samples. Underlying data are presented in Table 1.*

***Supplementary Table 4:*** *Differences between the popularity of responses to the question "How were you aware of transmission modelling?" across time periods within samples, according to Wilcoxon signed rank tests.* $p-values < 0.05$ *are considered statistically significant.*

| | | Number of respondents (n (%)) | | Wilcoxon signed rank test statistic | $n$ | $p-value$ |
|---|---|---|---|---|---|---|
| | | Prior to the COVID-19 pandemic | During the COVID-19 pandemic | | | |
| **Online panel** | **I was not aware** | 314 (62%) | 114 (23%) | 21318 | 504 | <0.001 |
| | **Newspaper (online or print)** | 69 (14%) | 204 (40%) | 288 | | <0.001 |
| | **News show (TV or online)** | 68 (13%) | 300 (60%) | 966 | | <0.001 |
| | **Social media** | 34 (7%) | 147 (29%) | 180 | | <0.001 |
| | **Internet search** | 49 (10%) | 122 (24%) | 588 | | <0.001 |
| | **Academic reports and papers** | 53 (11%) | 86 (17%) | 684 | | <0.001 |
| | **Other** | 36 (7%) | 19 (4%) | | | 0.005 |
| **Social media** | **I was not aware** | 74 (37%) | 6 (3%) | 2450 | 202 | <0.001 |
| | **Newspaper (online or print)** | 34 (17%) | 123 (61%) | 144 | | <0.001 |
| | **News show (TV or online)** | 27 (13%) | 142 (70%) | 183 | | <0.001 |
| | **Social media** | 16 (8%) | 125 (62%) | 56 | | <0.001 |
| | **Internet search** | 22 (11%) | 72 (36%) | 189 | | <0.001 |
| | **Academic reports and papers** | 48 (24%) | 97 (48%) | 244 | | <0.001 |
| | **Other** | 45 (22%) | 23 (11%) | 446 | | <0.001 |

***Supplementary Table 5:*** *Differences between the popularity of responses to the question "How were you aware of transmission modelling?" across samples within time periods, according to chi-squared tests.* $p - values < 0.05$ *are considered statistically significant.*

| | | Number of respondents (n (%)) | | $\chi^2_{df=1}$ | $n$ | $p - value$ |
|---|---|---|---|---|---|---|
| | | Online panel | Social media | | | |
| **Prior to the COVID-19 pandemic** | **I was not aware** | 314 (62%) | 74 (37%) | 37.35 | 706 | <0.001 |
| | **Newspaper (online or print)** | 69 (14%) | 34 (17%) | 0.90 | | 0.342 |
| | **News show (TV or online)** | 68 (13%) | 27 (13%) | <0.001 | | 1 |
| | **Social media** | 34 (7%) | 16 (8%) | 0.15 | | 0.698 |
| | **Internet search** | 49 (10%) | 22 (11%) | 0.11 | | 0.743 |
| | **Academic reports and papers** | 53 (11%) | 48 (24%) | 19.57 | | <0.001 |
| | **Other** | 36 (7%) | 45 (22%) | 31.05 | | <0.001 |
| **During the COVID-19 pandemic** | **I was not aware** | 114 (23%) | 6 (3%) | 38.09 | | <0.001 |
| | **Newspaper (online or print)** | 204 (40%) | 123 (61%) | 23.36 | | <0.001 |
| | **News show (TV or online)** | 300 (60%) | 142 (70%) | 6.70 | | 0.010 |
| | **Social media** | 147 (29%) | 125 (62%) | 63.79 | | <0.001 |
| | **Internet search** | 122 (24%) | 72 (36%) | 8.90 | | 0.003 |
| | **Academic reports and papers** | 86 (17%) | 97 (48%) | 70.37 | | <0.001 |
| | **Other** | 19 (4%) | 23 (11%) | 13.62 | | <0.001 |

### How much did you know about how transmission modelling has been used throughout the COVID-19 pandemic?

Supplementary Figure 2 presents responses to the question "How much did you know about how transmission modelling has been used throughout the COVID-19 pandemic?" under both the online panel and social media samples.

Supplementary Table 6 presents the results of the linear regression model regressing the answers to this question on age group and gender. The three responses provided as multiple-choice options, "Too little", "About right" and "Too much", were enumerated as -1, 0, 1, respectively.

Large numbers of respondents from both samples (46% from the online panel and 30% from social media) reported having "Too little" information about the use of transmission modelling during the pandemic, although this was significantly greater among the online panel (chi-squared test: $\chi^2_{df=1} = 15.55;\ n = 706;\ p - value < 0.001$). By comparison, social media respondents (8%) were significantly more likely to report having "Too much" information than the online panel (2%) ($\chi^2_{df=1} = 14.52;\ n = 706;\ p - value < 0.001$). However, it is important to remember that most of the social media sample recorded being aware of transmission modelling, either generally or specifically in policy, during the pandemic period. As mentioned in the main text, these differences could be driven by a combination of a significantly greater percentage of social respondents working in the "Education" and "Research" sectors and by their relationship to the accounts used to share the survey (Supplementary Table 1).

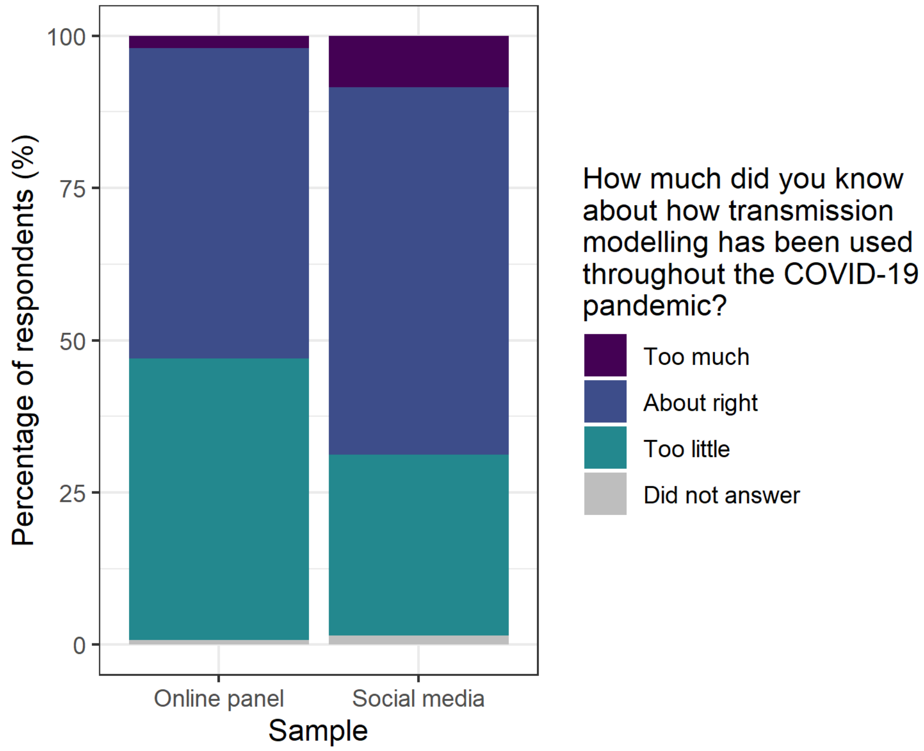

***Supplementary Figure 2:*** *Responses to the question "How much did you know about how transmission modelling has been used throughout the COVID-19 pandemic?" for the online panel and social media samples. Underlying data are presented in Table 1.*

***Supplementary Table 6:*** *Coefficient estimates, standard errors and p-values for the linear model regressing awareness of the use of transmission modelling during COVID-19 pandemic on age group (years) and gender within each model for each sample. Chi-squared tests for the overall significance of age and gender are also presented. The three responses provided as multiple-choice options, "Too little", "About right" and "Too much", were enumerated as -1, 0, 1, respectively.* $p - values < 0.05$ *are considered statistically significant.*

| **Sample** | **Response** | **Variable** | **Interpretation** | **Estimate** | **Standard Error** | **p-value** | **Chi-squared test** | |
|---|---|---|---|---|---|---|---|---|
| | | | | | | | $\chi^2$ | **p-value** |
| Online panel | How much do you know about how transmission modelling has been used throughout the COVID-19 pandemic? | Intercept | Baseline* | -0.52 | 0.07 | <0.001 | | |
| | | Age (years) | 26 – 35 | -0.00 | 0.09 | 0.999 | 1.20 (df=5) | 0.530 |
| | | | 36 – 45 | 0.06 | 0.09 | 0.508 | | |
| | | | 46 – 55 | 0.15 | 0.09 | 0.094 | | |
| | | | 56 – 65 | 0.04 | 0.08 | 0.655 | | |
| | | | 66+ | 0.08 | 0.11 | 0.444 | | |
| | | Gender | Male | 0.05 | 0.05 | 0.340 | 0.63 (df=1) | 0.535 |
| | | | Non-binary | 0.52 | 0.54 | 0.338 | | |
| | | | Prefer not to say | -0.18 | 0.31 | 0.567 | | |
| Social media | How much do you know about how transmission modelling has been used throughout the COVID-19 pandemic? | Intercept | Baseline* | -0.04 | 0.29 | 0.901 | | |
| | | Age (years) | 26 – 35 | 0.41 | 0.34 | 0.232 | 4.63 (df=5) | 0.018 |
| | | | 36 – 45 | -0.25 | 0.31 | 0.433 | | |
| | | | 46 – 55 | -0.17 | 0.30 | 0.560 | | |
| | | | 56 – 65 | -0.28 | 0.30 | 0.341 | | |
| | | | 66+ | -0.25 | 0.31 | 0.414 | | |
| | | Gender | Male | 0.05 | 0.08 | 0.555 | 0.12 (df=2) | 0.834 |
| | | | Prefer not to say | -0.01 | 0.34 | 0.966 | | |

*Baseline comprised of "Female" and "18 – 25" years.

### Level and means of awareness

Supplementary Figure 3 presents the results of “During the COVID-19 pandemic, how were you aware of transmission modelling?” stratified by responses to the question “How much do you know about how transmission modelling has been used throughout the COVID-19 pandemic?”. The underlying data are presented in Supplementary Table 7.

Almost all social media respondents selecting having “Too much” information on modelling during the pandemic, selected “Newspapers (online or print)” and “Academic reports and papers” as means of their awareness, with “Social media” also being a common response (Supplementary Table 7; Supplementary Figure 3). Within the online panel, those with “Too much” information were primarily aware through “News shows” (Supplementary Table 7; Supplementary Figure 3).

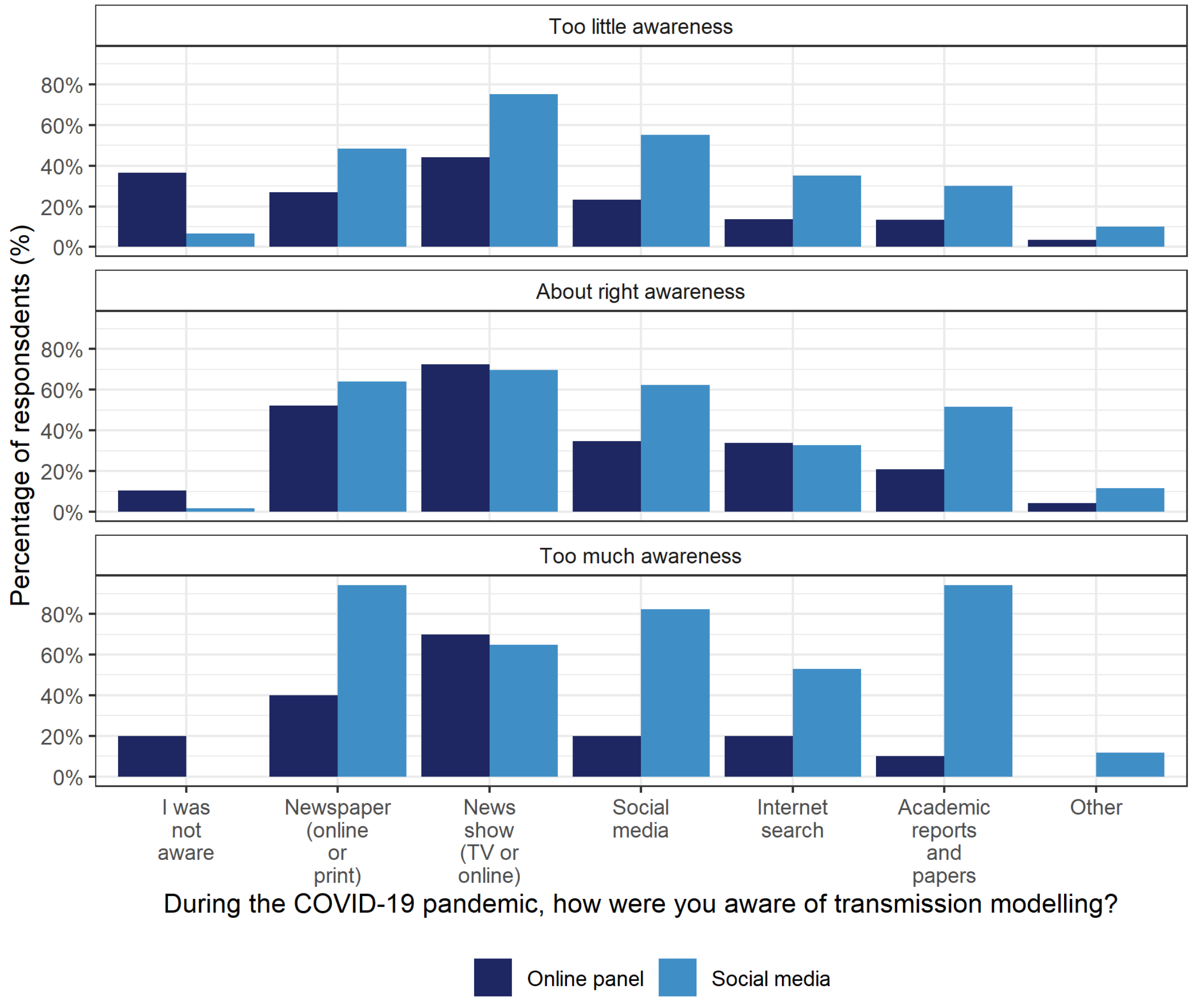

***Supplementary Figure 3:*** *Responses to the question “During the COVID-19 pandemic, how were you aware of transmission modelling?” stratified by responses to the question “How much did you know about how transmission modelling has been used throughout the COVID-19 pandemic?” for both the online panel and social media samples. Percentages are taken with respect to sample platform and level of awareness. Underlying data are presented in Supplementary Table 7.*

***Supplementary Table 7:*** *Number and percentage of responses under each category of the question "During the COVID-19 pandemic, how were you aware of transmission modelling?" stratified by responses to the question "How much did you know about how transmission modelling has been used throughout the COVID-19 pandemic?" for both the online panel and social media samples. As this question was open-ended, participants often provided more than one answer and so percentages do not represent mutually exclusive responses. Percentages taken with respect to the number selecting each awareness category within each sample as in Table 1.*

| | **Online panel** | | | **Social media** | | |
|---|---|---|---|---|---|---|
| | **Too little awareness** | **About right awareness** | **Too much awareness** | **Too little awareness** | **About right awareness** | **Too much awareness** |
| **I was not aware** | 85 (36%) | 27 (11%) | 2 (20%) | 4 (7%) | 2 (2%) | 0 (0%) |
| **Newspaper (online or print)** | 63 (27%) | 134 (52%) | 4 (40%) | 29 (48%) | 78 (64%) | 16 (94%) |
| **News show (TV or online)** | 103 (44%) | 186 (72%) | 7 (70%) | 45 (75%) | 85 (70%) | 11 (65%) |
| **Social media** | 54 (23%) | 89 (35%) | 2 (20%) | 33 (55%) | 76 (62%) | 14 (82%) |
| **Internet search** | 32 (14%) | 87 (34%) | 2 (20%) | 21 (35%) | 40 (33%) | 9 (53%) |
| **Academic reports and papers** | 31 (13%) | 54 (21%) | 1 (10%) | 18 (30%) | 63 (52%) | 16 (94%) |
| **Other** | 8 (3%) | 11 (4%) | 0 (0%) | 6 (10%) | 14 (11%) | 2 (12%) |

#### *Reliability of transmission modelling in informing public health policy*

On a scale of 1 – 10, with 1 being “extremely unreliable” and 10 being “extremely reliable” how did you feel about the use of transmission modelling in informing public health policy?

Supplementary Figure 4 presents histograms of the reliability scores within each sample across both time periods.

Supplementary Table 8 presents the results of the linear models regressing reliability score on age and gender, per sample and time period.

Supplementary Table 9 presents the results of the linear models regressing reliability score on age, gender, and awareness of the use of modelling in informing public health policy per sample and time period.

Supplementary Figure 5 and Supplementary Table 10 present the relationship between reliability scores prior to and during the COVID-19 pandemic under each sample.

Supplementary Table 11 presents the number and percentage of respondents within each sample changing reliability score in the period of the pandemic compared to the period prior.

Supplementary Table 12 presents a linear model regressing the difference in reliability scores on age group and gender. Difference in reliability score is defined as the score given during the pandemic minus the score given prior.

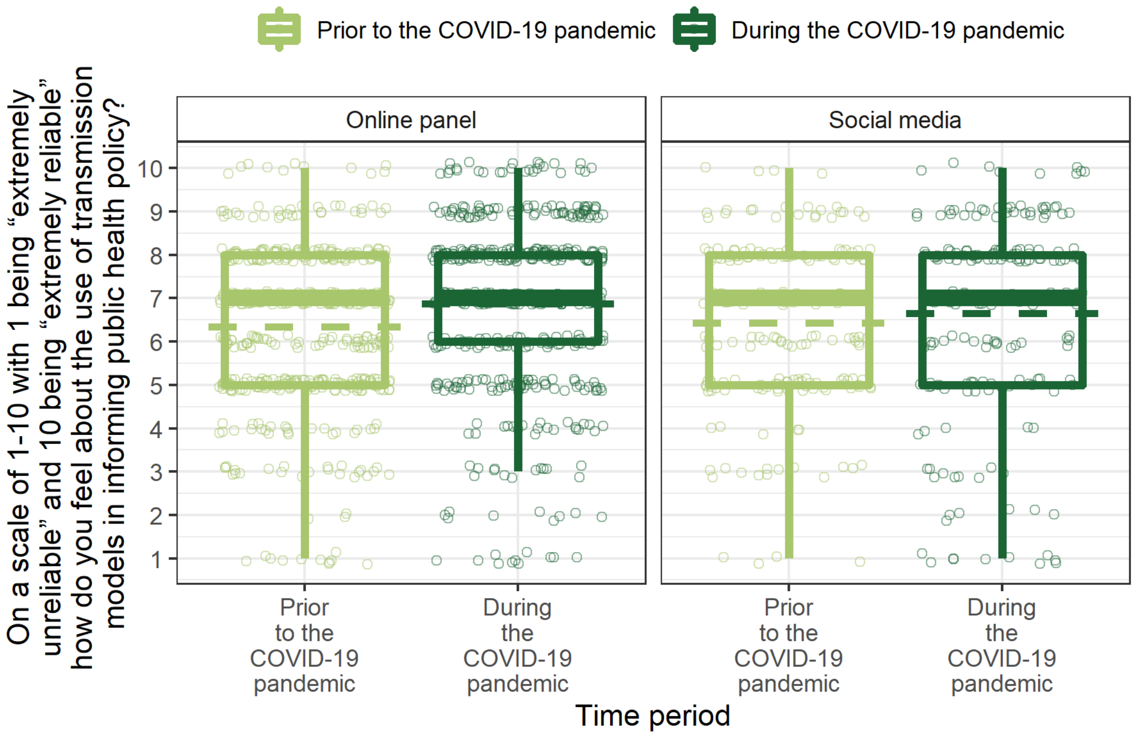

***Supplementary Figure 4:*** *Reliability of using transmission modelling to inform public health policy. (A) Responses to the question "On a scale of 1 – 10 with 1 being "extremely unreliable" and 10 being "extremely reliable" how do you feel about the use of transmission modelling in informing public health policy?" both "Prior to" and "During" the COVID19 pandemic, for both the online panel and social media samples. From bottom to top, the solid lines on the boxplot indicate: 1.5 times the interquartile range (IQR) less than the first quartile, first quartile, median, third quartile and 1.5 times the IQR greater than the third quartile. The dashed line corresponds to the mean. All responses are shown by the points, and so outliers, defined as any point outside the lower and upper bounds described, have been removed from the boxplots as they are shown in the presentation of the data. Points represent each reliability score and have been jittered to aid visual presentation. Underlying data are presented in Table 1.*

***Supplementary Table 8**: Coefficient estimates, standard errors and p-values for the linear regression models with age group and gender as predictors of reliability scores during each period and for each sample. Chi-squared tests for the overall significance of age and gender are also presented.* $p - values < 0.05$ *are considered statistically significant.*

| **Sample** | **Response** | **Variable** | **Interpretation** | **Estimate** | **Standard Error** | **p-value** | **Chi-squared test** | |
|---|---|---|---|---|---|---|---|---|
| | | | | | | | $\chi^2$ | **p-value** |
| Online panel | Reliability score prior to the COVID-19 pandemic | Intercept | Baseline* | 6.66 | 0.24 | <0.001 | | |
| | | Age (years) | 26 – 35 | -0.07 | 0.29 | 0.815 | 12.99 (df=5) | 0.567 |
| | | | 36 – 45 | -0.27 | 0.29 | 0.351 | | |
| | | | 46 – 55 | -0.48 | 0.29 | 0.106 | | |
| | | | 56 – 65 | -0.33 | 0.27 | 0.225 | | |
| | | | 66+ | -0.08 | 0.36 | 0.816 | | |
| | | Gender | Male | -0.20 | 0.17 | 0.234 | 10.19 (df=3) | 0.386 |
| | | | Non-binary | 0.41 | 1.84 | 0.824 | | |
| | | | Prefer not to say | 1.23 | 1.06 | 0.247 | | |
| Social media | Reliability score prior to the COVID-19 pandemic | Intercept | Baseline* | 6.95 | 0.96 | <0.001 | | |
| | | Age (years) | 26 – 35 | -0.89 | 1.12 | 0.426 | 4.66 (df=5) | 0.933 |
| | | | 36 – 45 | -0.36 | 1.03 | 0.728 | | |
| | | | 46 – 55 | -0.39 | 0.98 | 0.692 | | |
| | | | 56 – 65 | -0.24 | 0.97 | 0.807 | | |
| | | | 66+ | -0.60 | 1.02 | 0.554 | | |
| | | Gender | Male | -0.27 | 0.27 | 0.319 | 14.09 (df=2) | 0.141 |
| | | | Prefer not to say | -2.05 | 1.12 | 0.069 | | |
| Online panel | Reliability score during the COVID-19 pandemic | Intercept | Baseline* | 7.12 | 0.25 | <0.001 | | |
| | | Age (years) | 26 – 35 | -0.13 | 0.32 | 0.674 | 24.22 (df=5) | 0.290 |
| | | | 36 – 45 | -0.26 | 0.31 | 0.400 | | |
| | | | 46 – 55 | -0.63 | 0.32 | 0.049 | | |
| | | | 56 – 65 | -0.00 | 0.30 | 0.994 | | |
| | | | 66+ | -0.18 | 0.39 | 0.643 | | |
| | | Gender | Male | -0.11 | 0.18 | 0.526 | 3.18 (df=3) | 0.846 |
| | | | Non-binary | 0.01 | 1.99 | 0.994 | | |
| | | | Prefer not to say | 0.68 | 1.15 | 0.554 | | |
| Social media | Reliability score during the COVID-19 pandemic | Intercept | Baseline* | 7.84 | 1.11 | <0.001 | | |
| | | Age (years) | 26 – 35 | -2.87 | 1.29 | 0.027 | 59.66 (df=5) | 0.031 |
| | | | 36 – 45 | -1.28 | 1.18 | 0.281 | | |
| | | | 46 – 55 | -0.53 | 1.13 | 0.637 | | |
| | | | 56 – 65 | -0.58 | 1.12 | 0.607 | | |
| | | | 66+ | -0.62 | 1.17 | 0.598 | | |
| | | Gender | Male | -0.78 | 0.31 | 0.013 | 65.70 (df=2) | 0.001 |
| | | | Prefer not to say | -3.93 | 1.29 | 0.003 | | |

*Baseline comprised of “Female” and “18 – 25” years.

***Supplementary Table 9:*** *Coefficient estimates, standard errors and p-values for the linear regression models with age group, gender and awareness of the use of modelling in policy as predictors of reliability scores during each period and for each sample. The responses to the question "Were you aware of transmission modelling in informing public health policy?" used as an explanatory variable, "No", "Unsure" and "Yes" were enumerated as -1, 0, 1, respectively. Chi-squared tests for the overall significance of age and gender are also presented.* $p - values < 0.05$ *are considered statistically significant.*

| Sample | Response | Variable | Interpretation | Estimate | Standard Error | p-value | Chi-squared test | |
|---|---|---|---|---|---|---|---|---|
| | | | | | | | $\chi^2$ | p-value |
| Online panel | Reliability score prior to the COVID-19 pandemic | Intercept | Baseline* | 6.95 | 0.23 | <0.001 | | |
| | | Age (years) | 26 – 35 | -0.18 | 0.28 | 0.505 | 12.66 (df=5) | 0.520 |
| | | | 36 – 45 | -0.29 | 0.28 | 0.287 | | |
| | | | 46 – 55 | -0.62 | 0.28 | 0.028 | | |
| | | | 56 – 65 | -0.36 | 0.26 | 0.164 | | |
| | | | 66+ | 0.00 | 0.35 | 0.995 | | |
| | | Gender | Male | -0.31 | 0.16 | 0.050 | 10.65 (df=3) | 0.316 |
| | | | Non-binary | -0.92 | 1.75 | 0.597 | | |
| | | | Prefer not to say | 1.45 | 1.01 | 0.150 | | |
| | | Awareness | Transmission modelling used in policy | 0.69 | 0.09 | <0.001 | 174.06 (df=1) | <0.001 |
| Social media | Reliability score prior to the COVID-19 pandemic | Intercept | Baseline* | 6.85 | 0.96 | <0.001 | | |
| | | Age (years) | 26 – 35 | -0.94 | 1.10 | 0.394 | 4.66 (df=5) | 0.930 |
| | | | 36 – 45 | -0.38 | 1.01 | 0.706 | | |
| | | | 46 – 55 | -0.32 | 0.97 | 0.740 | | |
| | | | 56 – 65 | -0.19 | 0.96 | 0.841 | | |
| | | | 66+ | -0.59 | 1.01 | 0.558 | | |
| | | Gender | Male | -0.38 | 0.27 | 0.163 | 14.09 (df=2) | 0.135 |
| | | | Prefer not to say | -2.36 | 1.11 | 0.036 | | |
| | | Awareness | Transmission modelling used in policy | 0.37 | 0.16 | 0.024 | 17.99 (df=1) | 0.024 |
| Online panel | Reliability score during the COVID-19 pandemic | Intercept | Baseline* | 6.67 | 0.25 | <0.001 | | |
| | | Age (years) | 26 – 35 | -0.15 | 0.30 | 0.608 | 26.09 (df=5) | 0.192 |
| | | | 36 – 45 | -0.24 | 0.30 | 0.432 | | |
| | | | 46 – 55 | -0.83 | 0.30 | 0.007 | | |
| | | | 56 – 65 | -0.12 | 0.28 | 0.680 | | |
| | | | 66+ | -0.31 | 0.38 | 0.409 | | |
| | | Gender | Male | -0.14 | 0.17 | 0.424 | 3.07 (df=3) | 0.832 |
| | | | Non-binary | -0.40 | 1.89 | 0.832 | | |
| | | | Prefer not to say | 0.28 | 1.09 | 0.796 | | |
| | | Awareness | Transmission modelling used in policy | 0.88 | 0.12 | <0.001 | 205.75 (df=1) | <0.001 |
| Social media | Reliability score during the COVID-19 pandemic | Intercept | Baseline* | 7.93 | 1.26 | <0.001 | | |
| | | Age (years) | 26 – 35 | -2.89 | 1.28 | 0.026 | 55.94 (df=5) | 0.039 |
| | | | 36 – 45 | -1.05 | 1.18 | 0.374 | | |
| | | | 46 – 55 | -0.52 | 1.12 | 0.646 | | |
| | | | 56 – 65 | -0.56 | 1.11 | 0.613 | | |

| | | | | | | | | |
|---|---|---|---|---|---|---|---|---|
| | | | 66+ | -0.60 | 1.16 | 0.605 | | |
| | | Gender | Male | -0.71 | 0.31 | 0.025 | 61.25 (df=2) | 0.002 |
| | | | Prefer not to say | -3.89 | 1.28 | 0.003 | | |
| | | Awareness | Transmission modelling used in policy | -0.15 | 0.65 | 0.817 | 0.25 (df=1) | 0.817 |

*Baseline comprised of "Female" and "18 – 25" years.

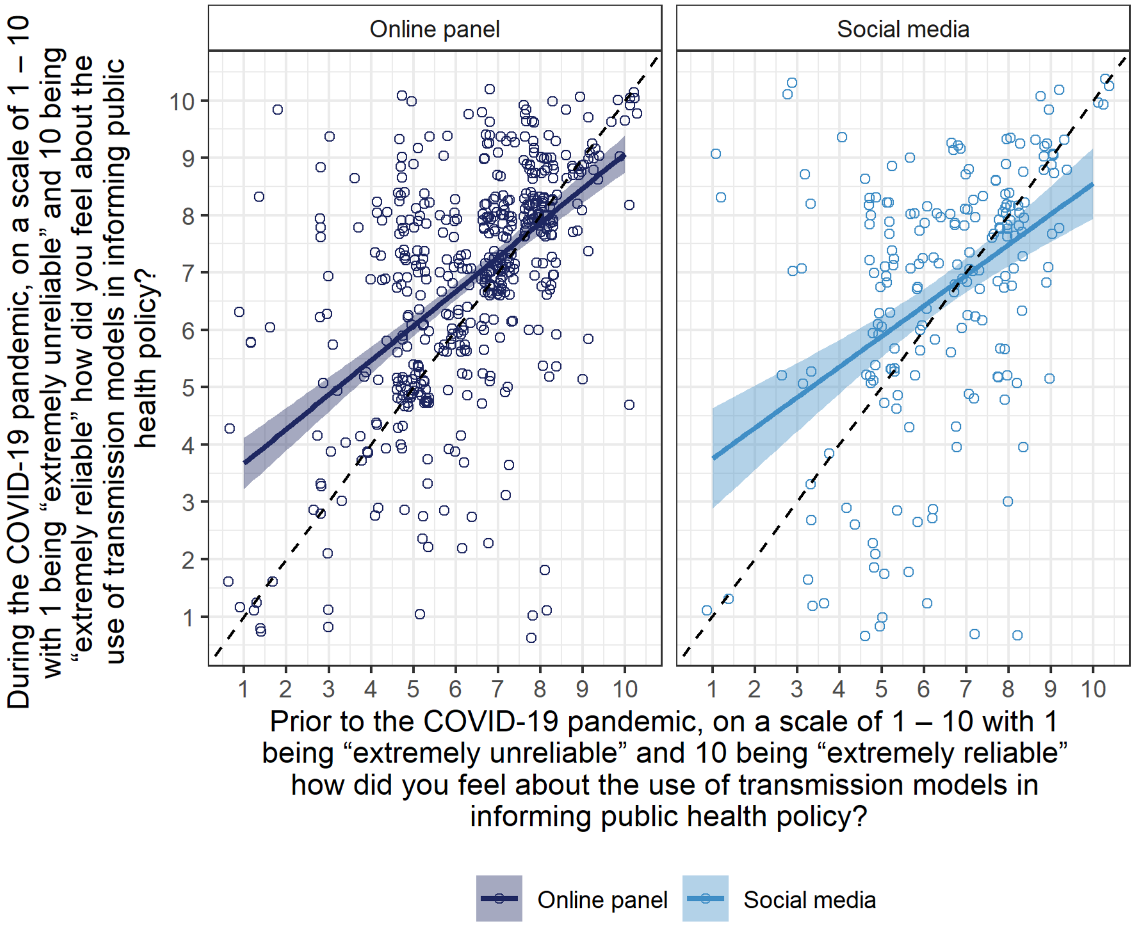

***Supplementary Figure 5:*** *Relationship between reliability scores given "Prior to" and "During" the COVID-19 pandemic for both the online panel and social media samples. Reliability scores obtained via the question: "On a scale of 1 – 10 with 1 being "extremely unreliable" and 10 being "extremely reliable" how did you feel about the use of transmission models in informing public health policy?". Points represent each reliability score and have been jittered to aid visual presentation. Coloured lines show the linear model presented in Supplementary Table 10, with shaded areas representing 95% confidence intervals. Black dashed line indicates where reliability scores during the pandemic equal those prior to the pandemic.*

***Supplementary Table 10:*** *Coefficient estimates, standard errors and p-values for the linear regression models with reliability score prior to the COVID-19 pandemic as the predictor of reliability score during the COVID-19 pandemic for each sample.*

| **Sample** | **Response** | **Variable** | **Estimate** | **Standard Error** | **p-value** |
|---|---|---|---|---|---|
| Online panel | Reliability score during the COVID-19 pandemic | Intercept | 3.07 | 0.27 | <0.001 |
| | | Reliability score prior to the COVID-19 pandemic | 0.60 | 0.04 | <0.001 |
| Social media | Reliability score during the COVID-19 pandemic | Intercept | 3.22 | 0.52 | <0.001 |
| | | Reliability score prior to the COVID-19 pandemic | 0.53 | 0.08 | <0.001 |

***Supplementary Table 11****: Number and percentage of respondents within each sample classified according to differences in reliability scores prior to and during the pandemic.*

| | | **Online panel** | **Social media** |
|---|---|---|---|
| **Number of respondents (n (%))** | **Reliability score increased** | 203 (40%) | 79 (39%) |
| | **Reliability score unchanged** | 217 (43%) | 70 (35%) |
| | **Reliability score decreased** | 76 (15%) | 52 (26%) |
| | **At least one score was missing** | 8 (2%) | 1 (0%) |

***Supplementary Table 12:*** *Coefficient estimates, standard errors and p-values for the linear regression models with age group and gender as predictors of the difference between reliability scores for each sample. Difference in reliability score is defined as the score given during the pandemic minus the score given prior. Chi-squared tests for the overall significance of age and gender are also presented.* $p - values < 0.05$ *are considered statistically significant.*

| Sample | Response | Variable | Interpretation | Estimate | Standard Error | p-value | Chi-squared test | |
|---|---|---|---|---|---|---|---|---|
| | | | | | | | $\chi^2$ | p-value |
| Online panel | Difference in reliability score | Intercept | Baseline* | 0.46 | 0.23 | 0.049 | | |
| | | Age (years) | 26 – 35 | -0.06 | 0.29 | 0.824 | 15.97 (df=5) | 0.420 |
| | | | 36 – 45 | 0.02 | 0.28 | 0.936 | | |
| | | | 46 – 55 | -0.19 | 0.29 | 0.520 | | |
| | | | 56 – 65 | 0.33 | 0.27 | 0.222 | | |
| | | | 66+ | -0.10 | 0.36 | 0.784 | | |
| | | Gender | Male | 0.09 | 0.16 | 0.576 | 2.24 (df=3) | 0.874 |
| | | | Non-binary | -0.39 | 1.80 | 0.828 | | |
| | | | Prefer not to say | -0.55 | 1.04 | 0.596 | | |
| Social media | Difference in reliability score | Intercept | Baseline* | 0.90 | 1.13 | 0.426 | | |
| | | Age (years) | 26 – 35 | -1.98 | 1.30 | 0.130 | 40.61 (df=5) | 0.142 |
| | | | 36 – 45 | -0.93 | 1.20 | 0.439 | | |
| | | | 46 – 55 | -0.15 | 1.14 | 0.895 | | |
| | | | 56 – 65 | -0.34 | 1.13 | 0.762 | | |
| | | | 66+ | -0.09 | 1.19 | 0.943 | | |
| | | Gender | Male | -0.53 | 0.32 | 0.095 | 21.16 (df=2) | 0.115 |
| | | | Prefer not to say | -1.89 | 1.31 | 0.150 | | |

*Baseline comprised of “Female” and “18 – 25” years.

### Reliability scores stratified by awareness

Supplementary Figure 6 presents reliability scores for both samples and time periods, stratified by responses to the question “Were you aware of transmission modelling being used in informing public health policy?”. There were statistically significant differences in reliability scores among those with awareness compared to those with no awareness among online panel respondents, holding time period constant, (Mann-Whitney U test: online panel and prior $p - value < 0.001$; online panel and during $p - value < 0.001$), but not among social media respondents (Mann-Whitney U test: social media and prior $p - value = 0.197$; social media and during $p - value = 0.148$).

Supplementary Figure 7 presents reliability scores according to responses to the question “How much do you know about how transmission modelling has been used throughout the COVID-19 pandemic?” for both samples. Online panel respondents who recorded an “About right” level of knowledge on the use of transmission modelling during the pandemic had significantly higher reliability scores than those stating they had “Too little” knowledge (Mann-Whitney U test: $p - value < 0.001$), but this was not a significant factor among social media respondents (Mann-Whitney U test: $p - value = 0.396$) (Supplementary Figure 7). The few respondents recording they had “Too much” knowledge had significantly lower reliability scores than those with “About right” knowledge within both samples (Mann-Whitney U test: Online panel $p - value = 0.002$; social media $p - value < 0.001$).

Supplementary Figure 8 presents reliability scores according to responses to the question “How were you aware of transmission modelling?” both “Prior to” and “During” the COVID-19 pandemic, for both samples. There was little distinction in reliability scores according to the means by which respondents were aware of modelling, with scores distributed across the entire scale under each option within each sample.

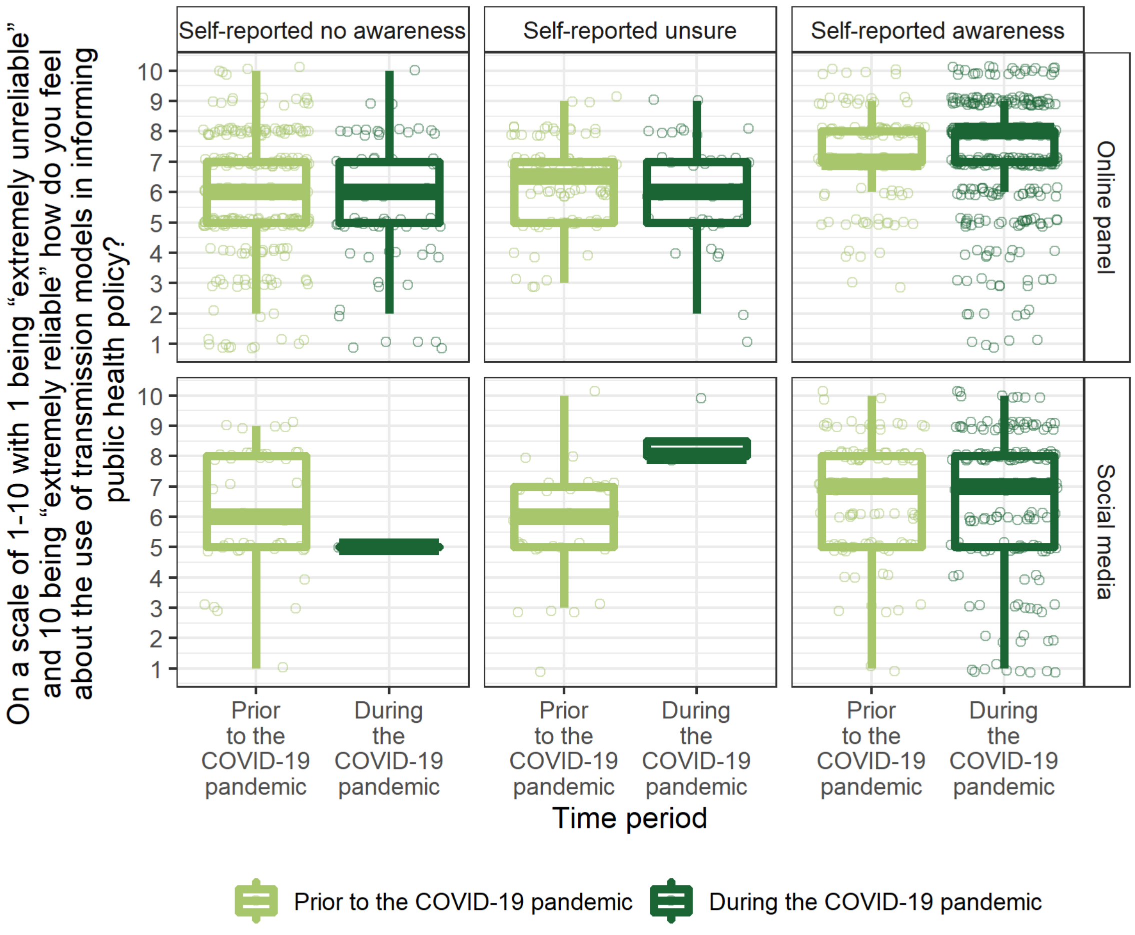

***Supplementary Figure 6:*** *Responses to the question "On a scale of 1 – 10 with 1 being "extremely unreliable" and 10 being "extremely reliable" how do you feel about the use of transmission modelling in informing public health policy?" both "Prior to" and "During" the COVID-19 pandemic, for both the online panel and social media samples, stratified by self-reported level of awareness according to the question "Were you aware of the use of transmission models in informing public health policy?" both "Prior to" and "During" the COVID-19 pandemic. From bottom to top, the solid lines on the boxplot indicate: 1.5 times the interquartile range (IQR) less than the first quartile, first quartile, median, third quartile and 1.5 times the IQR greater than the third quartile. All responses are shown by the points, and so outliers, defined as any point outside the lower and upper bounds described, have been removed from the boxplots as they are shown in the presentation of the data. Points represent each reliability score and have been jittered to aid visual presentation.*

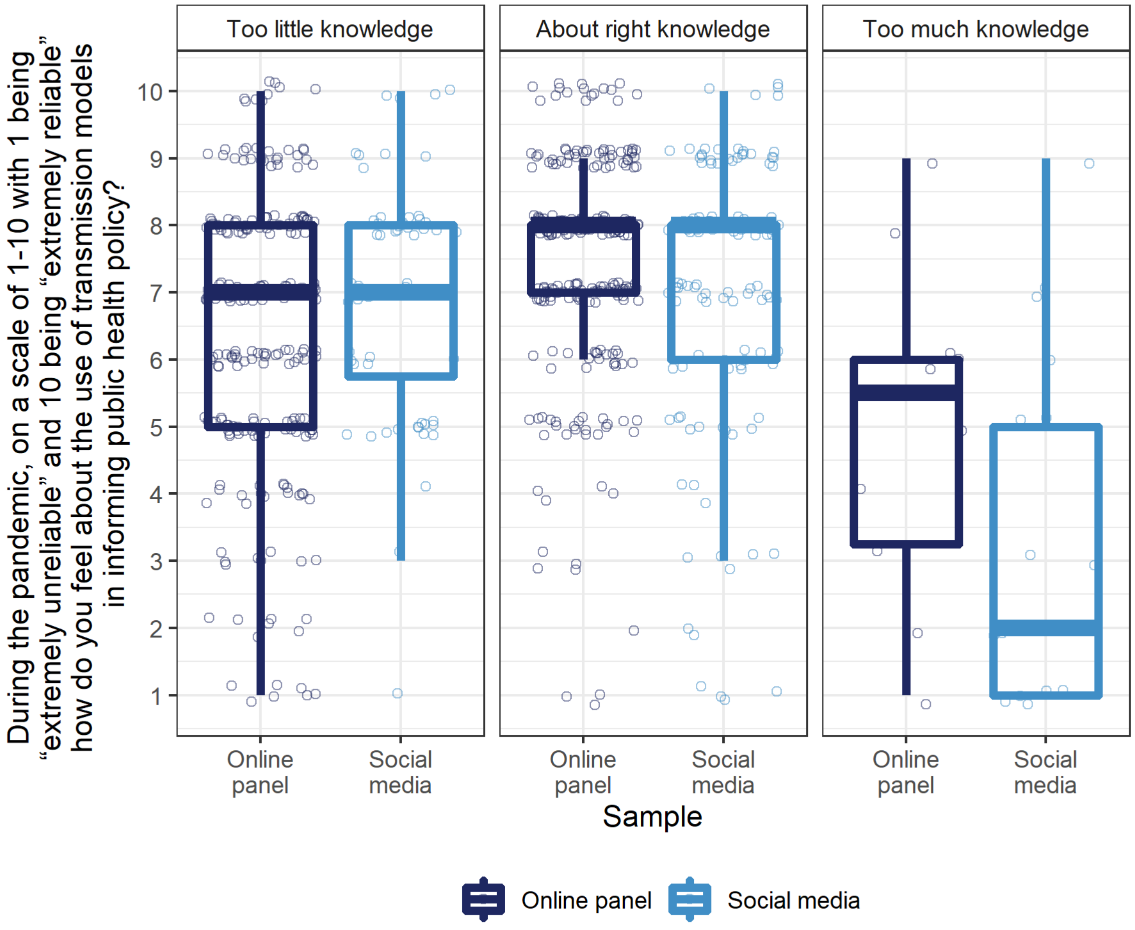

***Supplementary Figure 7:*** *Responses to the question "During the COVID-19 pandemic, on a scale of 1 – 10 with 1 being "extremely unreliable" and 10 being "extremely reliable" how do you feel about the use of transmission modelling in informing public health policy?" for both the online panel and social media samples, stratified by self-reported level of awareness according to the question "How much do you know about how transmission modelling has been used throughout the COVID-19 pandemic?". From bottom to top, the solid lines on the boxplot indicate: 1.5 times the interquartile range (IQR) less than the first quartile, first quartile, median, third quartile and 1.5 times the IQR greater than the third quartile. The dashed line corresponds to the mean. All responses are shown by the points, and so outliers, defined as any point outside the lower and upper bounds described, have been removed from the boxplots as they are shown in the presentation of the data. Points represent each reliability score and have been jittered to aid visual presentation.*

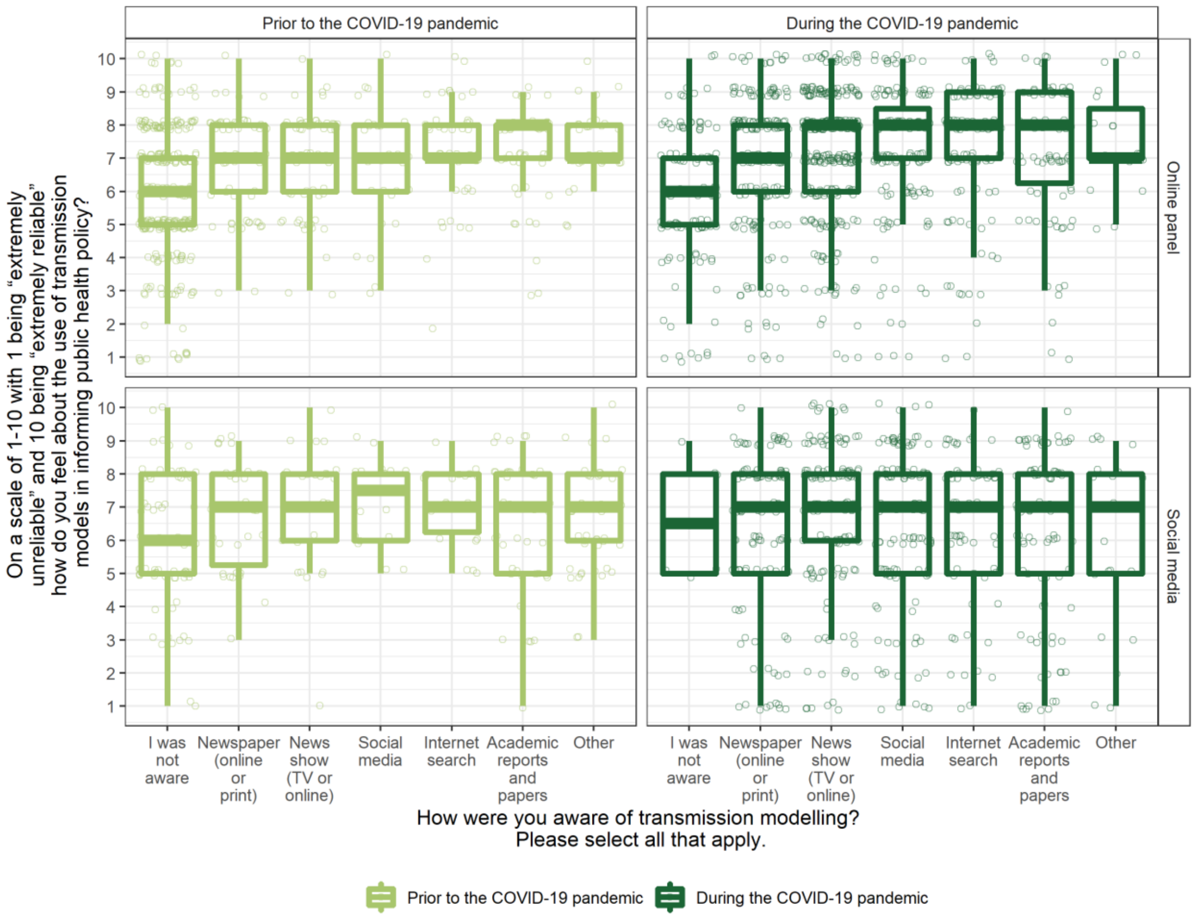

***Supplementary Figure 8:*** *Responses to the question "On a scale of 1 – 10 with 1 being "extremely unreliable" and 10 being "extremely reliable" how do you feel about the use of transmission modelling in informing public health policy?" both "Prior to" and "During" the COVID-19 pandemic, for both the online panel and social media samples, stratified by responses to the question "How were you aware of transmission modelling?" both "Prior to" and "During" the COVID-19 pandemic. From bottom to top, the solid lines on the boxplot indicate: 1.5 times the interquartile range (IQR) less than the first quartile, first quartile, median, third quartile and 1.5 times the IQR greater than the third quartile. The dashed line corresponds to the mean. All responses are shown by the points, and so outliers, defined as any point outside the lower and upper bounds described, have been removed from the boxplots as they are shown in the presentation of the data. Points represent each reliability score and have been jittered to aid visual presentation.*

*Trust in government advice*

#### How much did you trust government advice regarding public health issues?

Supplementary Figure 9 presents reliability scores stratified by answers to question "How much did you trust government advice regarding public health issues?" both "Prior to" and "During" the COVID-19 pandemic for both samples. Supplementary Table 13 present the median and variances of the reliability scores for the same stratifications.

Supplementary Table 14 presents the number and percentage of respondents within each sample classified according to differences in responses to the aforementioned question across time periods.

Supplementary Table 15 presents the results of the corresponding linear model regressing the responses to the question "How much did you trust government advice regarding public health issues?" both "Prior to" and "During" the COVID-19 pandemic on age group and gender of respondents within each sample. The three levels of trust provided in the multiple-choice options "No level of trust whatsoever", "Moderate level of trust" and "High level of trust" were enumerated as -1, 0, 1 respectively.

Supplementary Table 16 presents the results of the linear model regressing changes in responses to the question "How much did you trust government advice regarding public health issues?" in the period of the pandemic compared to the period prior, on age group and gender within each sample. The three responses provided in the multiple-choice options, "No trust whatsoever", "Moderate level of trust" and "High level of trust", were enumerated as -1, 0, 1, respectively, and so the differences (response) could take values of -2, -1, 0, 1, 2.

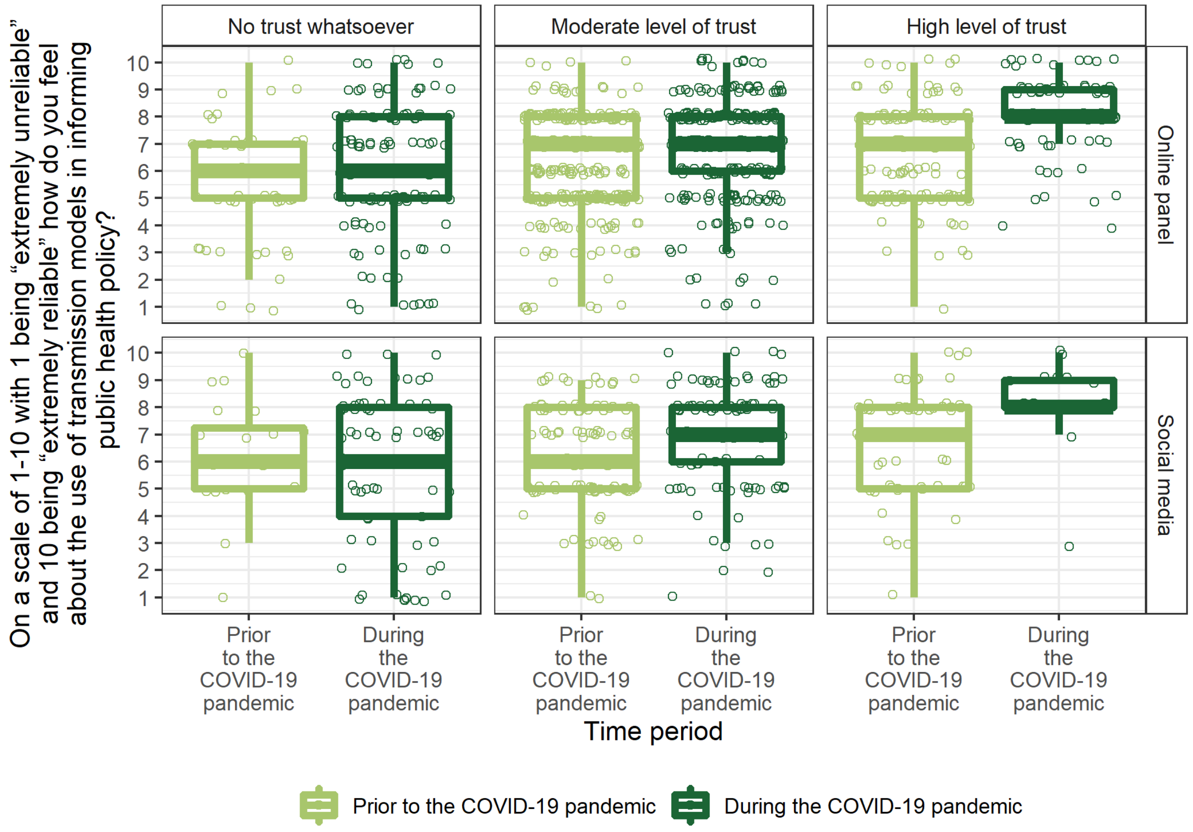

***Supplementary Figure 9:*** *Responses to the question "On a scale of 1 – 10 with 1 being "extremely unreliable" and 10 being "extremely reliable" how do you feel about the use of transmission modelling in informing public health policy?" both "Prior to" and "During" the COVID-19 pandemic, for both the online panel and social media samples, stratified by responses to the question "How much did you trust government advice regarding public health issues?" both "Prior to" and "During" the COVID-19 pandemic. From bottom to top, the solid lines on the boxplot indicate: 1.5 times the interquartile range (IQR) less than the first quartile, first quartile, median, third quartile and 1.5 times the IQR greater than the third quartile. The dashed line corresponds to the mean. All responses are shown by the points, and so outliers, defined as any point outside the lower and upper bounds described, have been removed from the boxplots as they are shown in the presentation of the data. Points represent each reliability score and have been jittered to aid visual presentation.*

***Supplementary Table 13:*** *Median and variances of the responses to the question question "On a scale of 1 – 10 with 1 being "extremely unreliable" and 10 being "extremely reliable" how do you feel about the use of transmission modelling in informing public health policy?" for both the online panel and social media samples stratified by responses to the question "How much did you trust government advice regarding public health issues?" both "Prior to" and "During" the COVID-19 pandemic.*

| | | No trust whatsoever | | Moderate level of trust | | High level of trust | |
|---|---|---|---|---|---|---|---|
| | | Median | Variance | Median | Variance | Median | Variance |
| Online panel | Prior to the COVID-19 pandemic | 6.00 | 4.61 | 7.00 | 3.06 | 7.00 | 3.21 |
| | During the COVID-19 pandemic | 6.00 | 5.35 | 7.00 | 3.21 | 8.00 | 1.83 |
| Social media | Prior to the COVID-19 pandemic | 6.00 | 4.41 | 6.00 | 3.35 | 7.00 | 3.47 |
| | During the COVID-19 pandemic | 6.00 | 7.09 | 7.00 | 3.50 | 8.00 | 2.26 |

***Supplementary Table 14****: Number and percentage of respondents within each sample classified according to differences in responses to the question "How much did you trust government advice regarding public health issues?" both "Prior to" and "During" the COVID-19 pandemic.*

| | | **Online panel** | **Social media** |
|---|---|---|---|
| **Number of respondents (n (%))** | **Trust in government advice increased** | 40 (8%) | 12 (6%) |
| | **Trust in government advice unchanged** | 328 (65%) | 92 (46%) |
| | **Trust in government advice decreased** | 135 (27%) | 98 (49%) |
| | **At least one score was missing** | 1 (0%) | 0 (0%) |

***Supplementary Table 15:*** *Coefficient estimates, standard errors and p-values for the linear model regressing responses to the question "How much did you trust government advice regarding public health issues?" on age group and gender during each period and for each sample. The three responses provided in the multiple-choice options, "No trust whatsoever", "Moderate level of trust" and "High level of trust", were enumerated as -1, 0, 1, respectively. Chi-squared tests for the overall significance of age and gender are also presented.* $p - values < 0.05$ *are considered statistically significant.*

| Sample | Response | Variable | Interpretation | Estimate | Standard Error | p-value | Chi-squared test | |
|---|---|---|---|---|---|---|---|---|
| | | | | | | | $\chi^2$ | p-value |
| Online panel | Trust in government prior to the COVID-19 pandemic | Intercept | Baseline* | 0.11 | 0.07 | 0.136 | | |
| | | Age (years) | 26 – 35 | 0.02 | 0.09 | 0.831 | 0.13 (df=5) | 0.995 |
| | | | 36 – 45 | 0.01 | 0.09 | 0.953 | | |
| | | | 46 – 55 | -0.01 | 0.09 | 0.933 | | |
| | | | 56 – 65 | -0.02 | 0.09 | 0.795 | | |
| | | | 66+ | -0.02 | 0.12 | 0.886 | | |
| | | Gender | Male | 0.10 | 0.05 | 0.047 | 1.46 (df=3) | 0.230 |
| | | | Non-binary | -0.13 | 0.59 | 0.822 | | |
| | | | Prefer not to say | 0.22 | 0.34 | 0.515 | | |
| Social media | Trust in government prior to the COVID-19 pandemic | Intercept | Baseline* | 0.23 | 0.31 | 0.461 | | |
| | | Age (years) | 26 – 35 | 0.25 | 0.36 | 0.485 | 2.25 (df=5) | 0.312 |
| | | | 36 – 45 | -0.07 | 0.33 | 0.838 | | |
| | | | 46 – 55 | -0.00 | 0.32 | 0.998 | | |
| | | | 56 – 65 | 0.01 | 0.31 | 0.969 | | |
| | | | 66+ | -0.24 | 0.33 | 0.465 | | |
| | | Gender | Male | 0.02 | 0.09 | 0.776 | 0.05 (df=2) | 0.939 |
| | | | Prefer not to say | 0.09 | 0.36 | 0.805 | | |
| Online panel | Trust in government during the COVID-19 pandemic | Intercept | Baseline* | -0.17 | 0.08 | 0.039 | | |
| | | Age (years) | 26 – 35 | -0.02 | 0.10 | 0.864 | 1.96 (df=5) | 0.455 |
| | | | 36 – 45 | 0.11 | 0.10 | 0.279 | | |
| | | | 46 – 55 | 0.05 | 0.10 | 0.664 | | |
| | | | 56 – 65 | 0.12 | 0.10 | 0.233 | | |
| | | | 66+ | 0.15 | 0.13 | 0.238 | | |
| | | Gender | Male | 0.08 | 0.06 | 0.167 | 1.71 (df=3) | 0.251 |
| | | | Non-binary | -0.81 | 0.65 | 0.213 | | |
| | | | Prefer not to say | -0.23 | 0.38 | 0.539 | | |
| Social media | Trust in government during the COVID-19 pandemic | Intercept | Baseline* | -0.48 | 0.31 | 0.122 | | |
| | | Age (years) | 26 – 35 | -0.00 | 0.36 | 0.993 | 2.90 (df=5) | 0.175 |
| | | | 36 – 45 | 0.02 | 0.33 | 0.964 | | |
| | | | 46 – 55 | 0.18 | 0.32 | 0.564 | | |
| | | | 56 – 65 | 0.37 | 0.31 | 0.243 | | |
| | | | 66+ | 0.15 | 0.33 | 0.656 | | |
| | | Gender | Male | -0.02 | 0.09 | 0.811 | 2.22 (df=2) | 0.053 |
| | | | Prefer not to say | -0.88 | 0.36 | 0.016 | | |

*Baseline comprised of "Female" and "18 – 25" years.

***Supplementary Table 16:*** *Coefficient estimates, standard errors and p-values for the linear model regressing changes in responses to the question "How much did you trust government advice regarding public health issues?" in the period of the pandemic compared to the period prior, on age group and gender within each sample. The three responses provided in the multiple-choice options, "No trust whatsoever", "Moderate level of trust" and "High level of trust", were enumerated as -1, 0, 1, respectively, and so the differences (response) could take integer values between -2 and 2. Chi-squared tests for the overall significance of age and gender are also presented.* $p-values < 0.05$ *are considered statistically significant.*

| Sample | Response | Variable | Interpretation | Estimate | Standard Error | p-value | Chi-squared test | |
|---|---|---|---|---|---|---|---|---|
| | | | | | | | $\chi^2$ | p-value |
| Online panel | Change in trust in government regarding public health issues | Intercept | Baseline* | -0.30 | 0.09 | <0.001 | | |
| | | Age (years) | 26 – 35 | -0.02 | 0.11 | 0.845 | 2.81 (df=5) | 0.269 |
| | | | 36 – 45 | 0.12 | 0.10 | 0.242 | | |
| | | | 46 – 55 | 0.07 | 0.11 | 0.511 | | |
| | | | 56 – 65 | 0.15 | 0.10 | 0.118 | | |
| | | | 66+ | 0.19 | 0.13 | 0.158 | | |
| | | Gender | Male | -0.03 | 0.06 | 0.634 | 1.11 (df=3) | 0.469 |
| | | | Non-binary | -0.68 | 0.67 | 0.307 | | |
| | | | Prefer not to say | -0.45 | 0.38 | 0.239 | | |
| Social media | Change in trust in government regarding public health issues | Intercept | Baseline* | -0.72 | 0.37 | 0.054 | | |
| | | Age (years) | 26 – 35 | -0.26 | 0.43 | 0.549 | 4.56 (df=5) | 0.126 |
| | | | 36 – 45 | 0.08 | 0.39 | 0.832 | | |
| | | | 46 – 55 | 0.18 | 0.37 | 0.625 | | |
| | | | 56 – 65 | 0.35 | 0.37 | 0.341 | | |
| | | | 66+ | 0.39 | 0.38 | 0.319 | | |
| | | Gender | Male | -0.05 | 0.10 | 0.657 | 2.70 (df=2) | 0.078 |
| | | | Prefer not to say | -0.97 | 0.43 | 0.024 | | |

*Baseline comprised of "Female" and "18 – 25" years.

### Trust in government advice and awareness

Supplementary Table 17 presents the results of the linear model regressing responses to the question “How much did you trust government advice regarding public health issues?” on age group, gender and awareness of the use of modelling in policy during each period and for each sample. The three responses provided in the multiple-choice options for the response, “No trust whatsoever”, “Moderate level of trust” and “High level of trust”, were enumerated as -1, 0, 1, respectively. Similarly, the responses to the question “Were you aware of transmission modelling in informing public health policy?” used as an explanatory variable, “No”, “Unsure” and “Yes” were enumerated as -1, 0, 1, respectively. Data to the response question stratified by awareness are presented in Supplementary Figure 10, with the percentages underlying this figure set out in Supplementary Table 18.

***Supplementary Table 17:*** *Coefficient estimates, standard errors and p-values for the linear model regressing responses to the question "How much did you trust government advice regarding public health issues?" on age group, gender and awareness of the use of modelling in policy during each period and for each sample. The three responses provided in the multiple-choice options for the response, "No trust whatsoever", "Moderate level of trust" and "High level of trust", were enumerated as -1, 0, 1, respectively. Similarly, the responses to the question "Were you aware of transmission modelling in informing public health policy?" used as an explanatory variable, "No", "Unsure" and "Yes" were enumerated as -1, 0, 1, respectively. Chi-squared tests for the overall significance of explanatory variables are also presented.* $p - values < 0.05$ *are considered statistically significant.*

| Sample | Response | Variable | Interpretation | Estimate | Standard Error | p-value | Chi-squared test | |
|---|---|---|---|---|---|---|---|---|
| | | | | | | | $\chi^2$ | p-value |
| Online panel | Trust in government prior to the COVID-19 pandemic | Intercept | Baseline* | 0.11 | 0.08 | 0.149 | | |
| | | Age (years) | 26 – 35 | 0.02 | 0.09 | 0.828 | 0.18 (df=5) | 0.991 |
| | | | 36 – 45 | 0.01 | 0.09 | 0.942 | | |
| | | | 46 – 55 | -0.01 | 0.09 | 0.937 | | |
| | | | 56 – 65 | -0.03 | 0.09 | 0.734 | | |
| | | | 66+ | -0.02 | 0.12 | 0.886 | | |
| | | Gender | Male | 0.11 | 0.05 | 0.042 | 1.53 (df=3) | 0.212 |
| | | | Non-binary | -0.13 | 0.59 | 0.824 | | |
| | | | Prefer not to say | 0.22 | 0.34 | 0.510 | | |
| | | Awareness | Transmission modelling used in policy | -0.00 | 0.03 | 0.979 | 0 (df=1) | 0.979 |
| Social media | Trust in government prior to the COVID-19 pandemic | Intercept | Baseline* | 0.25 | 0.31 | 0.420 | | |
| | | Age (years) | 26 – 35 | 0.26 | 0.36 | 0.466 | 2.45 (df=5) | 0.309 |
| | | | 36 – 45 | -0.06 | 0.33 | 0.849 | | |
| | | | 46 – 55 | -0.01 | 0.32 | 0.964 | | |
| | | | 56 – 65 | 0.00 | 0.31 | 0.992 | | |
| | | | 66+ | -0.25 | 0.33 | 0.450 | | |
| | | Gender | Male | 0.05 | 0.09 | 0.605 | 0.048 (df=2) | 0.938 |
| | | | Prefer not to say | 0.15 | 0.36 | 0.678 | | |
| | | Awareness | Transmission modelling used in policy | -0.07 | 0.05 | 0.159 | 0.75 (df=1) | 0.159 |
| Online panel | Trust in government during the COVID-19 pandemic | Intercept | Baseline* | -0.20 | 0.09 | 0.022 | | |
| | | Age (years) | 26 – 35 | -0.01 | 0.10 | 0.938 | 1.66 (df=5) | 0.551 |
| | | | 36 – 45 | 0.11 | 0.10 | 0.294 | | |
| | | | 46 – 55 | 0.02 | 0.10 | 0.829 | | |
| | | | 56 – 65 | 0.11 | 0.10 | 0.258 | | |
| | | | 66+ | 0.12 | 0.13 | 0.344 | | |
| | | Gender | Male | 0.08 | 0.06 | 0.185 | 1.70 (df=3) | 0.254 |
| | | | Non-binary | -0.84 | 0.65 | 0.195 | | |
| | | | Prefer not to say | -0.26 | 0.38 | 0.497 | | |
| | | Awareness | Transmission modelling used in policy | 0.05 | 0.04 | 0.209 | 0.66 (df=1) | 0.209 |

| | | | | | | | | |
|---|---|---|---|---|---|---|---|---|
| Social media | Trust in government during the COVID-19 pandemic | Intercept | Baseline* | -0.41 | 0.36 | 0.256 | | |
| | | Age (years) | 26 – 35 | -0.02 | 0.36 | 0.960 | 2.67 (df=5) | 0.217 |
| | | | 36 – 45 | 0.03 | 0.33 | 0.917 | | |
| | | | 46 – 55 | 0.18 | 0.32 | 0.563 | | |
| | | | 56 – 65 | 0.37 | 0.31 | 0.242 | | |
| | | | 66+ | 0.15 | 0.33 | 0.654 | | |
| | | Gender | Male | -0.01 | 0.09 | 0.929 | 2.21 (df=2) | 0.055 |
| | | | Prefer not to say | -0.87 | 0.36 | 0.017 | | |
| | | Awareness | Transmission modelling used in policy | -0.09 | 0.18 | 0.641 | 0.08 (df=1) | 0.641 |

*Baseline comprised of “Female” and “18 – 25” years.

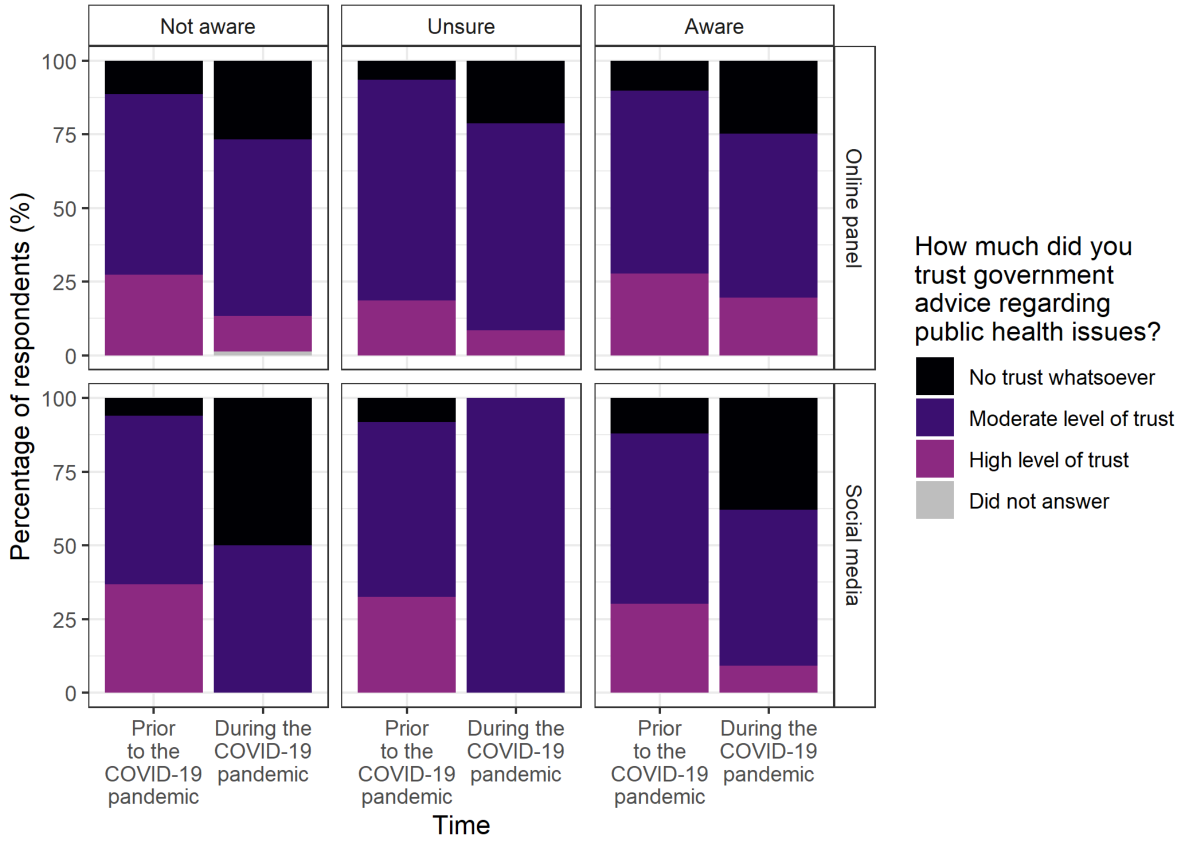

***Supplementary Figure 10:*** *The percentage of observations from both the online panel and social samples according to time period and answer to the question “How much did you trust government advice regarding public health issues?“ both “Prior to” and “During” the COVID-19 pandemic, stratified by responses to the question “Were you aware of the use of transmission models in informing public health policy?” both “Prior to” and “During” the COVID-19 pandemic. Underlying data are presented in Supplementary Table 18.*

***Supplementary Table 18:*** *The percentage of observations from both the online panel and social media samples according to time period and answer to the question "How much did you trust government advice regarding public health issues?" both "Prior to" and "During" the COVID-19 pandemic, stratified by responses to the question "Were you aware of the use of transmission models in informing public health policy?" both "Prior to" and "During" the COVID-19 pandemic.*

| **Platform** | **Time** | **Were you aware of the use of transmission models in informing public health policy?** | **How much did you trust government advice regarding public health issues?** | | | |
|---|---|---|---|---|---|---|
| | | | **High level of trust** | **Moderate level of trust** | **No trust whatsoever** | **Did not answer** |
| Online panel | Prior to the COVID-19 pandemic | Yes | 38 (28%) | 85 (62%) | 14 (10%) | 0 (0%) |
| | | Unsure | 17 (19%) | 68 (75%) | 6 (7%) | 0 (0%) |
| | | No | 74 (27%) | 168 (61%) | 31 (11%) | 0 (0%) |
| | During the COVID-19 pandemic | Yes | 73 (20%) | 208 (56%) | 93 (25%) | 0 (0%) |
| | | Unsure | 4 (9%) | 33 (70%) | 10 (21%) | 0 (0%) |
| | | No | 9 (12%) | 45 (60%) | 20 (27%) | 1 (1%) |
| Social media | Prior to the COVID-19 pandemic | Yes | 35 (30%) | 67 (58%) | 14 (12%) | 0 (0%) |
| | | Unsure | 12 (32%) | 22 (59%) | 3 (8%) | 0 (0%) |
| | | No | 18 (37%) | 28 (57%) | 3 (6%) | 0 (0%) |
| | During the COVID-19 pandemic | Yes | 18 (9%) | 103 (53%) | 74 (38%) | 0 (0%) |
| | | Unsure | 0 (0%) | 4 (100%) | 0 (0%) | 0 (0%) |
| | | No | 0 (0%) | 1 (50%) | 1 (50%) | 0 (0%) |

#### How do you feel when government advice changes based on new scientific evidence?

Supplementary Table 19 presents the results of the corresponding linear model regressing the responses to the question “How do you feel when government advice changes based on new scientific evidence?” on gender and age group of respondents within each sample. The three levels of trust provided in the multiple-choice options “I have less trust in the advice”, “My level of trust remains unchanged” and “I have more trust in the advice” were enumerated as -1, 0, and 1, respectively.

Supplementary Table 20 expands the regression presented in Supplementary Table 19 but also controls for responses to the question “Were you aware of the use of transmission models in informing public health policy?” both “Prior to” and “During” the COVID-19 pandemic. Responses to this question, “Yes”, “Unsure” and “No” were enumerated as 1, 0, and -1, respectively.

Analogously to Supplementary Table 20, Supplementary Table 21 expands the regression in Supplementary Table 19 but also controls for responses to the question “How much did you trust government scientific advice regarding public health policy?” both “Prior to” and “During” the COVID-19 pandemic. Responses to this question, “High level of trust”, “Moderate level of trust” and “No trust whatsoever” were enumerated as 1, 0, and -1, respectively.

***Supplementary Table 19:*** *Coefficient estimates, standard errors and p-values for the linear model regressing responses to the question "How do you feel when government advice changes based on new scientific evidence?" on age group and gender for each sample. The three responses provided in the multiple-choice options for the response, "I have less trust in the advice", "My level of trust remains unchanged" and "I have more trust in the advice", were enumerated as -1, 0, 1, respectively. Chi-squared tests for the overall significance of age and gender are also presented.* $p - values < 0.05$ *are considered statistically significant.*

| Sample | Response | Variable | Interpretation | Estimate | Standard Error | p-value | Chi-squared test | |
|---|---|---|---|---|---|---|---|---|
| | | | | | | | $\chi^2$ | p-value |
| Online panel | Feeling regarding government changing advice based on new scientific evidence | Intercept | Baseline* | 0.43 | 0.09 | <0.001 | | |
| | | Age (years) | 26 – 35 | -0.06 | 0.11 | 0.560 | 2.91 (df=5) | 0.304 |
| | | | 36 – 45 | -0.12 | 0.11 | 0.255 | | |
| | | | 46 – 55 | -0.24 | 0.11 | 0.033 | | |
| | | | 56 – 65 | -0.16 | 0.10 | 0.126 | | |
| | | | 66+ | -0.21 | 0.14 | 0.137 | | |
| | | Gender | Male | 0.01 | 0.06 | 0.898 | 0.50 (df=3) | 0.790 |
| | | | Non-binary | -0.37 | 0.70 | 0.599 | | |
| | | | Prefer not to say | 0.35 | 0.40 | 0.386 | | |
| Social media | Feeling regarding government changing advice based on new scientific evidence | Intercept | Baseline* | 0.60 | 0.34 | 0.075 | | |
| | | Age (years) | 26 – 35 | 0.08 | 0.39 | 0.839 | 1.39 (df=5) | 0.669 |
| | | | 36 – 45 | -0.18 | 0.36 | 0.611 | | |
| | | | 46 – 55 | 0.02 | 0.34 | 0.957 | | |
| | | | 56 – 65 | -0.07 | 0.34 | 0.834 | | |
| | | | 66+ | -0.20 | 0.36 | 0.576 | | |
| | | Gender | Male | -0.14 | 0.09 | 0.147 | 2.75 (df=2) | 0.045 |
| | | | Prefer not to say | -0.87 | 0.39 | 0.028 | | |

*Baseline comprised of "Female" and "18 – 25" years.

***Supplementary Table 20:*** *Coefficient estimates, standard errors and p-values for the generalised linear model regressing responses to the question "How do you feel when government advice changes based on new scientific evidence?" on age group and gender for each sample. The three responses provided in the multiple-choice options for the response, "I have less trust in the advice", "My level of trust remains unchanged" and "I have more trust in the advice", were enumerated as -1, 0, 1, respectively. Responses to the explanatory variable regarding awareness, "Yes", "Unsure" and "No", were enumerated as 1, 0, -1, respectively. Chi-squared tests for the overall significance of explanatory are also presented.* $p - values < 0.05$ *are considered statistically significant.*

| Sample | Response | Variable | Interpretation | Estimate | Standard Error | p-value | Chi-squared test $\chi^2$ | Chi-squared test p-value |
|---|---|---|---|---|---|---|---|---|
| Online panel | Feeling regarding government changing advice based on new scientific evidence | Intercept | Baseline* | 0.41 | 0.09 | <0.001 | | |
| | | Age (years) | 26 – 35 | -0.08 | 0.11 | 0.470 | 2.55 (df=5) | 0.372 |
| | | | 36 – 45 | -0.14 | 0.11 | 0.208 | | |
| | | | 46 – 55 | -0.26 | 0.11 | 0.020 | | |
| | | | 56 – 65 | -0.16 | 0.10 | 0.118 | | |
| | | | 66+ | -0.18 | 0.14 | 0.202 | | |
| | | Gender | Male | 0.01 | 0.06 | 0.932 | 0.52 (df=3) | 0.776 |
| | | | Non-binary | -0.36 | 0.70 | 0.604 | | |
| | | | Prefer not to say | 0.34 | 0.40 | 0.403 | | |
| | | Awareness | Transmission modelling used in policy prior to the pandemic | 0.05 | 0.04 | 0.187 | 1.85 (df=1) | 0.049 |
| | | | Transmission modelling used in policy during the pandemic | 0.08 | 0.04 | 0.061 | 1.68 (df=1) | 0.060 |
| Social media | Feeling regarding government changing advice based on new scientific evidence | Intercept | Baseline* | 0.57 | 0.39 | 0.142 | | |
| | | Age (years) | 26 – 35 | 0.08 | 0.39 | 0.840 | 1.20 (df=5) | 0.734 |
| | | | 36 – 45 | -0.12 | 0.36 | 0.747 | | |
| | | | 46 – 55 | 0.03 | 0.34 | 0.929 | | |
| | | | 56 – 65 | -0.06 | 0.34 | 0.853 | | |
| | | | 66+ | -0.19 | 0.35 | 0.593 | | |
| | | Gender | Male | -0.13 | 0.10 | 0.173 | 2.53 (df=2) | 0.056 |
| | | | Prefer not to say | -0.89 | 0.39 | 0.025 | | |
| | | Awareness | Transmission modelling used in policy prior to the pandemic | 0.04 | 0.06 | 0.534 | 0.18 (df=1) | 0.515 |
| | | | Transmission modelling used in policy during the pandemic | 0.01 | 0.20 | 0.945 | 0.00 (df=1) | 0.945 |

*Baseline comprised of "Female" and "18 – 25" years.

***Supplementary Table 21:*** *Coefficient estimates, standard errors and p-values for the generalised linear model regressing responses to the question "How do you feel when government advice changes based on new scientific evidence?" on age group and gender for each sample. The three responses provided in the multiple-choice options for the response, "I have less trust in the advice", "My level of trust remains unchanged" and "I have more trust in the advice", were enumerated as -1, 0, 1, respectively. Responses to the explanatory variable regarding government trust, "High level of trust", "Moderate level of trust" and "No trust whatsoever", were enumerated as 1, 0, -1, respectively. Chi-squared tests for the overall significance of age and gender are also presented.* $p-values < 0.05$ *are considered statistically significant.*

| Sample | Response | Variable | Interpretation | Estimate | Standard Error | p-value | Chi-squared test | |
|---|---|---|---|---|---|---|---|---|
| | | | | | | | $\chi^2$ | p-value |
| Online panel | Feeling regarding government changing advice based on new scientific evidence | Intercept | Baseline* | 0.44 | 0.09 | <0.001 | | |
| | | Age (years) | 26 – 35 | -0.05 | 0.11 | 0.624 | 2.74 (df=5) | 0.323 |
| | | | 36 – 45 | -0.13 | 0.11 | 0.237 | | |
| | | | 46 – 55 | -0.23 | 0.11 | 0.036 | | |
| | | | 56 – 65 | -0.16 | 0.10 | 0.116 | | |
| | | | 66+ | -0.21 | 0.14 | 0.121 | | |
| | | Gender | Male | -0.01 | 0.06 | 0.889 | 0.50 (df=3) | 0.788 |
| | | | Non-binary | -0.27 | 0.70 | 0.702 | | |
| | | | Prefer not to say | 0.37 | 0.40 | 0.362 | | |
| | | Government trust | Trust in advice prior to the pandemic | 0.05 | 0.06 | 0.446 | 1.70 (df=1) | 0.059 |
| | | | Trust in advice during the pandemic | 0.12 | 0.05 | 0.024 | 2.42 (df=1) | 0.024 |
| Social media | Feeling regarding government changing advice based on new scientific evidence | Intercept | Baseline* | 0.65 | 0.34 | 0.057 | | |
| | | Age (years) | 26 – 35 | 0.06 | 0.39 | 0.877 | 1.39 (df=5) | 0.662 |
| | | | 36 – 45 | -0.18 | 0.36 | 0.620 | | |
| | | | 46 – 55 | -0.01 | 0.34 | 0.984 | | |
| | | | 56 – 65 | -0.12 | 0.34 | 0.723 | | |
| | | | 66+ | -0.20 | 0.35 | 0.574 | | |
| | | Gender | Male | -0.14 | 0.09 | 0.152 | 2.75 (df=2) | 0.043 |
| | | | Prefer not to say | -0.76 | 0.40 | 0.057 | | |
| | | Government trust | Trust in advice prior to the pandemic | 0.08 | 0.08 | 0.340 | 0.99 (df=1) | 0.131 |
| | | | Trust in advice during the pandemic | 0.13 | 0.08 | 0.108 | 1.12 (df=1) | 0.108 |

*Baseline comprised of "Female" and "18 – 25" years.

### Trust in changing advice and awareness

Supplementary Figure 11 presents reliability scores for both samples and time periods stratified by responses to the question “How do you feel when government advice changes based on new scientific evidence?”.

Supplementary Figure 12 presents the results of the question “How do you feel when government advice changes based on new scientific evidence?” stratified by response to the question “How much did you trust government advice regarding public health issues?” both “Prior to” and “During” the COVID-19 pandemic and under both samples. Supplementary Table 22 presents the data underlying this figure.

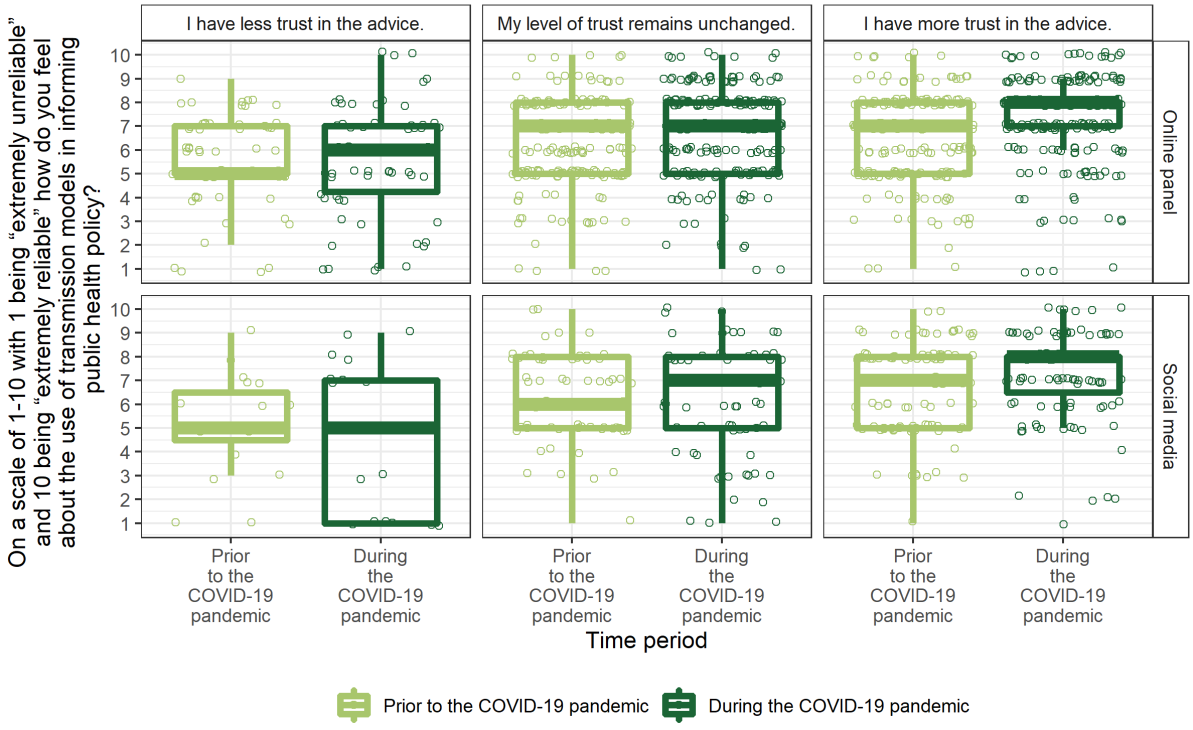

***Supplementary Figure 11:*** *Responses to the question “On a scale of 1 – 10 with 1 being “extremely unreliable” and 10 being “extremely reliable” how do you feel about the use of transmission modelling in informing public health policy?” both “Prior to” and “During” the COVID-19 pandemic, for both the online panel and social media samples, stratified by responses to the question “How much do you feel when government advice changes based on new scientific evidence?”. From bottom to top, the solid lines on the boxplot indicate: 1.5 times the interquartile range (IQR) less than the first quartile, first quartile, median, third quartile and 1.5 times the IQR greater than the third quartile. The dashed line corresponds to the mean. All responses are shown by the points, and so outliers, defined as any point outside the lower and upper bounds described, have been removed from the boxplots as they are shown in the presentation of the data. Points represent each reliability score and have been jittered to aid visual presentation.*

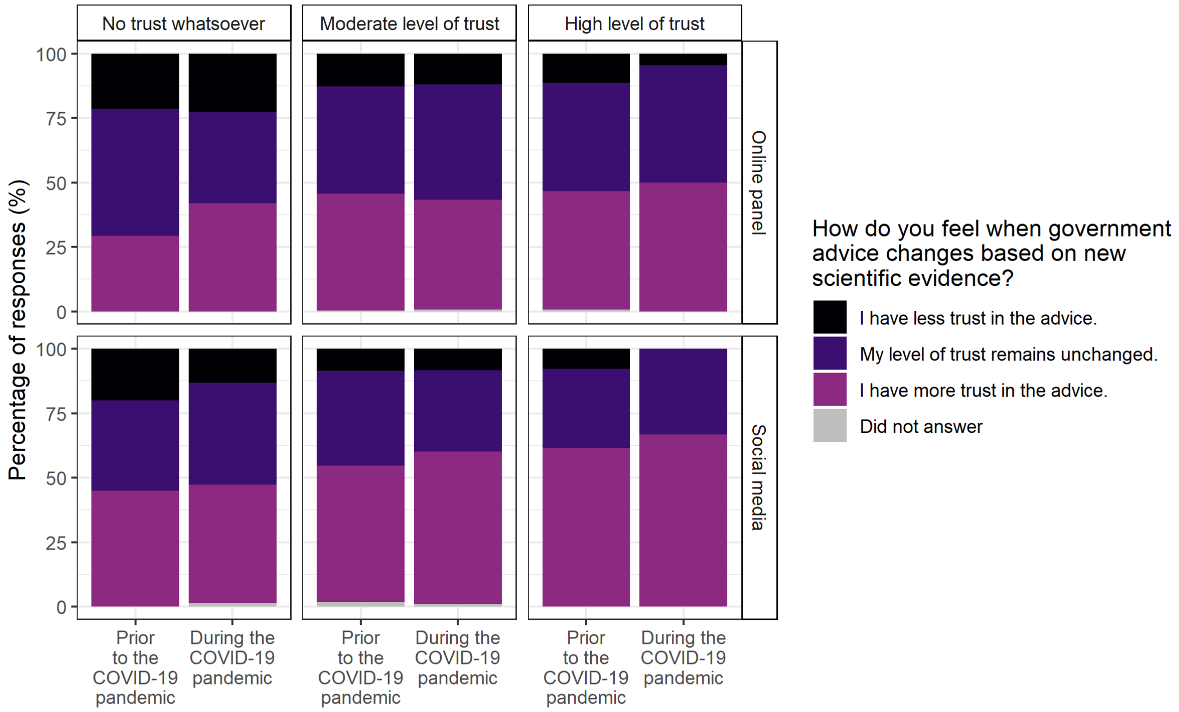

***Supplementary Figure 12:*** *Responses to the question "How do you feel when government advice changes based on new scientific evidence?" for both the online panel and social media samples, stratified by answers to the question "How much did you trust government advice regarding public health policy?" at the time "Prior to" and "During" the COVID-19 pandemic. Underlying data are presented in Supplementary Table 22.*

***Supplementary Table 22:*** *The percentage of observations from both the online panel and social media samples according to time period and answer to the question "How do you feel when government advice changes based on new scientific evidence?", stratified by responses to the question "Were you aware of the use of transmission models in informing public health policy?" both "Prior to" and "During" the COVID-19 pandemic.*

| **Platform** | **Time** | **How much did you trust government advice regarding public health policy?** | **How do you feel when government advice changes based on new scientific evidence?** | | | |
|---|---|---|---|---|---|---|
| | | | **I have more trust in the advice.** | **My level of trust remains unchanged.** | **I have less trust in the advice.** | **Did not answer** |
| Online panel | Prior to the COVID-19 pandemic | High level of trust | 60 (46%) | 55 (42%) | 15 (11%) | 1 (1%) |
| | | Moderate level of trust | 146 (45%) | 134 (42%) | 41 (13%) | 1 (0%) |
| | | No trust whatsoever | 15 (29%) | 25 (49%) | 11 (22%) | 0 (0%) |
| | During the COVID-19 pandemic | High level of trust | 44 (50%) | 40 (45%) | 4 (5%) | 0 (0%) |
| | | Moderate level of trust | 124 (43%) | 130 (45%) | 35 (12%) | 2 (1%) |
| | | No trust whatsoever | 52 (42%) | 44 (35%) | 28 (23%) | 0 (0%) |
| Social media | Prior to the COVID-19 pandemic | High level of trust | 40 (62%) | 20 (31%) | 5 (8%) | 0 (0%) |
| | | Moderate level of trust | 62 (53%) | 43 (37%) | 10 (9%) | 2 (2%) |
| | | No trust whatsoever | 9 (45%) | 7 (35%) | 4 (20%) | 0 (0%) |
| | During the COVID-19 pandemic | High level of trust | 12 (67%) | 6 (33%) | 0 (0%) | 0 (0%) |
| | | Moderate level of trust | 64 (59%) | 34 (31%) | 9 (8%) | 1 (1%) |
| | | No trust whatsoever | 35 (46%) | 30 (39%) | 10 (13%) | 1 (1%) |

*Responsibility of communicating modelling to the public*

Supplementary Figure 13 presents the responses to the question "Who do you think has the responsibility of ensuring that the public are informed about the use of modelling in policy decisions, particularly in the COVID-19 pandemic?" for both samples.

Supplementary Table 23 presents the results of chi-squared tests for statistically significant differences in the proportion of respondents selecting multiple-choice answers to the question "Who do you think has the responsibility of ensuring that the public are informed about the use of modelling in policy decisions, particularly in the COVID-19 pandemic?" between the two samples.

Supplementary Figure 14 presents the results to the aforementioned question stratified by answers to the question "How much did you know about how transmission modelling has been used throughout the COVID-19 pandemic?". The robustness of these results to awareness suggests that this is not an influencing factor.

Of the few who selected "None of the above" to the aforementioned question, of which there were 1 from the online panel and 2 from the social media sample, respondents were either unsure (online panel), believed in a "collaborative" (social media) approach or the "NHS" (social media).

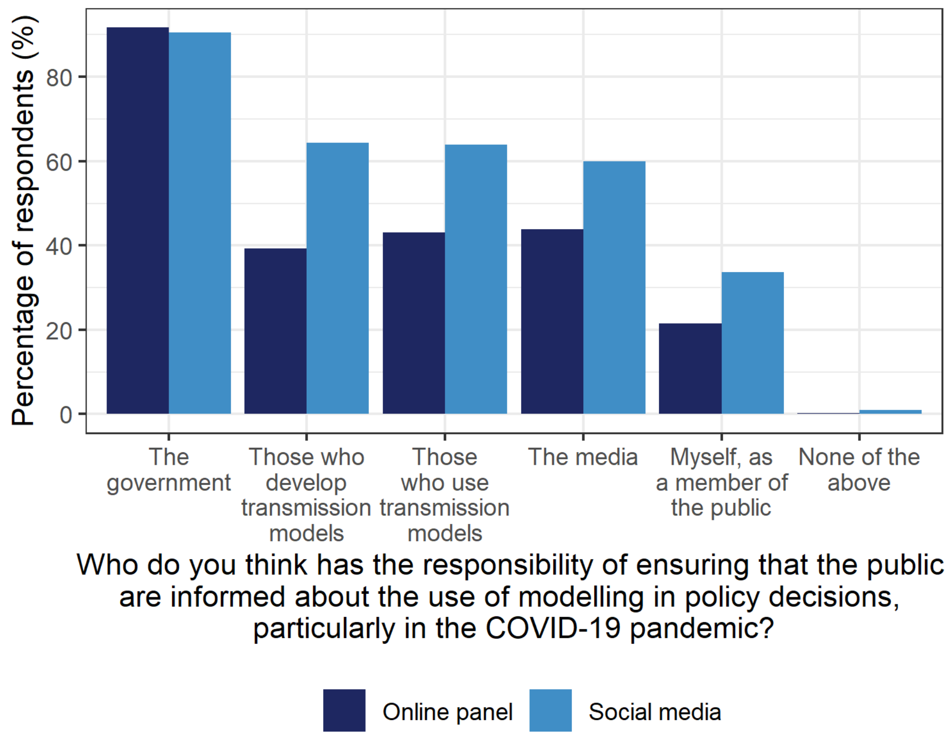

***Supplementary Figure 13:*** *Responses to the question "Who do you think has the responsibility of ensuring that the public are informed about the use of modelling in policy decisions, particularly in the COVID-19 pandemic? Please select all that apply." for both samples. Underlying data are presented in Table 1.*

***Supplementary Table 23:*** *Results of chi-squared tests for statistically significant differences in the proportion of respondents selecting multiple-choice answers to the question "Who do you think has the responsibility of ensuring that the public are informed about the use of modelling in policy decisions, particularly in the COVID-19 pandemic?" between the online panel and social media samples. Note that respondents could select multiple responses for this question.* $p - values < 0.05$ *are considered statistically significant.*

| | **Number of respondents (n (%))** | | $\chi^2$ | $n$ | $p - value$ |
|---|---|---|---|---|---|
| | **Online panel** | **Social media** | | | |
| **The government** | 462 (92%) | 183 (91%) | 0.10 | 706 | 0.756 |
| **Those who develop transmission models** | 198 (39%) | 130 (64%) | 35.44 | | <0.001 |
| **Those who use transmission models** | 217 (43%) | 129 (64%) | 24.15 | | <0.001 |
| **The media** | 221 (44%) | 121 (60%) | 14.24 | | <0.001 |
| **Myself, as a member of the public** | 108 (21%) | 68 (34%) | 10.89 | | <0.001 |
| **None of the above** | 1 (0%) | 2 (1%) | 0.67 | | 0.411 |

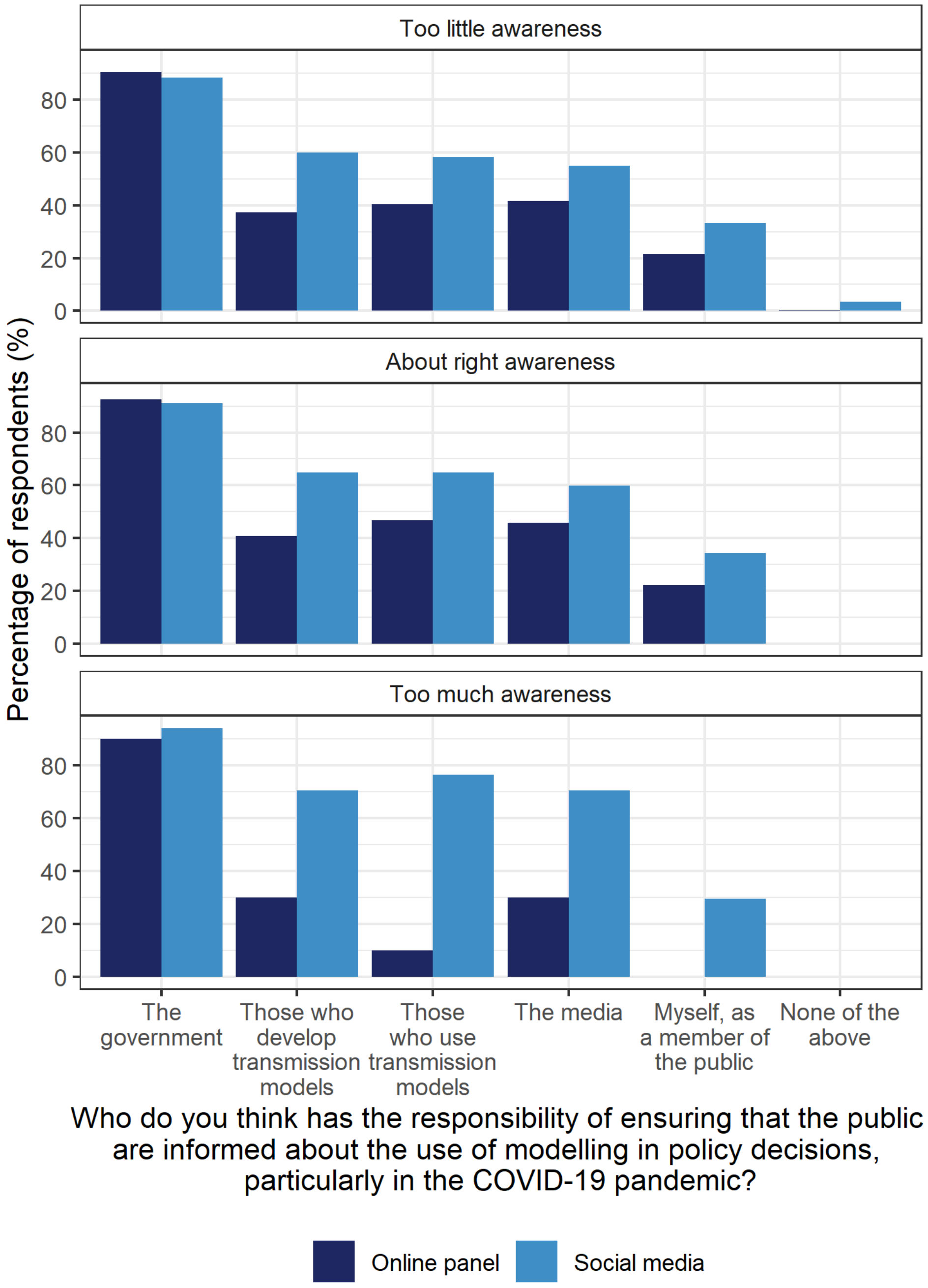

***Supplementary Figure 14**: Responses to the question "Who do you think has the responsibility of ensuring that the public are informed about the use of modelling in policy decisions, particularly in the COVID-19 pandemic? Please select all that apply." for both the online panel and social media samples, stratified by answers to the question "How much do you know about how transmission modelling has been used throughout the COVID-19 pandemic?".*

#### *Where do those who develop transmission models work?*

Supplementary Figure 15 presents responses to the question "Where do you think those who developed and used transmission models work?" both "Prior to" and "During" the COVID-19 pandemic for both samples. The underlying data are presented in Supplementary Table 24, with Wilcoxon signed rank tests for significant differences across time presented in Supplementary Table 25. For each category, data were enumerated to a binary scale with 1 indicating that this category was referred to and 0 indicating that it was not. Supplementary Table 26 presents the results of chi-squared tests for differences in the proportion of individual selecting each option across samples.

Many respondents from the online panel stated that they considered those who developed transmission models worked either within unspecified research institutions (35%) or within academia specifically (33%) prior to the COVID-19 pandemic. "Research"-related answers remained the most common responses in the period of the pandemic. However, significantly more responses referred to "Academia" specifically (36%; $p - value = 0.027$; Supplementary Table 25) compared to research within unspecified institutions which decreased in popularity (30%; $p - value = 0.021$; Supplementary Table 25). The number of respondents who believed that modellers worked within "Government" during the pandemic rose significantly (29% during up from 18% prior; Supplementary Table 25), as did "Healthcare services" (14% during up from 8% prior; Supplementary Table 25). In particular, direct mention of the NHS doubled during (6%) compared to prior (3%) to the pandemic (Supplementary Table 25).

Within the social media sample, there were few categories with statistically significant differences in the percentage of respondents selecting them across the two time periods (Supplementary Table 25). "Research (Academia)" was the most common response across both time periods (70% prior and 73% during; Supplementary Table 25). The second most common response was "Public health bodies" (28% in both time periods), which was similarly popular among this cohort as it was among online panel respondents in each time period. However, "Research (unspecified affiliations)" was a significantly less common response among social media respondents in both time periods compared to the online panel sample (Supplementary Table 26). "Healthcare services" was also a comparatively less popular choice among this cohort than online panel respondents among both time periods, as was explicit mention of the NHS in the period of the pandemic but not in the period prior (Supplementary Table 26).

In both samples, respondents referred to "Advisory roles" specifically in the period during the pandemic (4% online panel and 7% social media, respectively; Supplementary Table 26), but not at all in the period prior. Social media respondents were borderline significantly more likely to select "Advisory roles" than online panel respondents (Supplementary Table 26). Furthermore, "SAGE" was explicitly mentioned in the period of the pandemic, with approximately equal percentages of respondents selecting this option under both samples.

In the period prior to the pandemic, social media respondents were equally likely as online panel respondents to record an answer related to "Government". However, in the period of the pandemic, social media respondents were borderline significantly less likely to answer related to "Government" than online panel respondents (Supplementary Table 26). Although "Research (Academia)" was significantly more common among both samples than "Government" in the period of the pandemic (Wilcoxon signed rank tests: online panel: $p - value = 0.016$; social media:$p - value < 0.001$), the magnitude of the differences was substantially different (online panel: 36% "Research (Academia)" vs. 29% "Government"; social media: 73% "Research (Academia)" vs. 22% "Government").

The percentage of respondents answering “Pharmaceutical companies” was relatively small and remained constant across samples and time periods. This was similar when considering “Media”. As observed with other questions, significantly more respondents in the online panel sample answered “Unsure” in both time periods compared to the social sample (online panel: 8% prior and 5% during; social media: 1% prior and 0% during; Supplementary Figure 15; Supplementary Table 24).

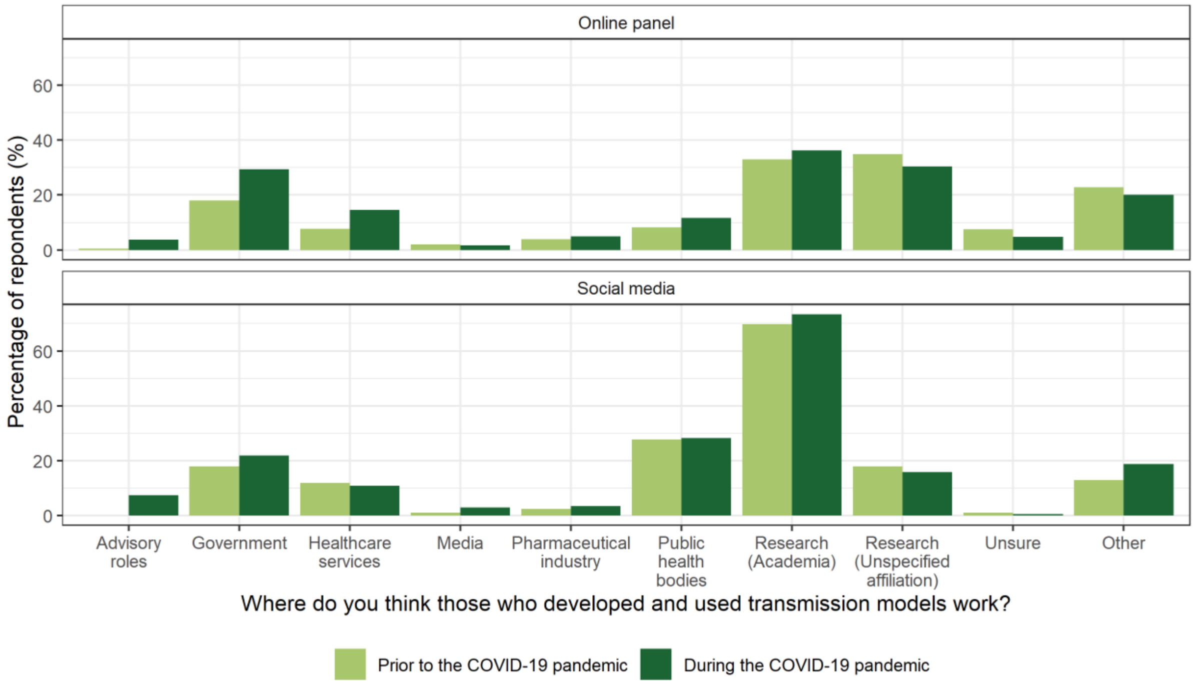

***Supplementary Figure 15:*** *Responses to the question “Where do you think those who developed and used transmission models work?” either “Prior to” or “During” the COVID-19 pandemic for both the online panel and social media respondents.*

***Supplementary Table 24:*** *Responses to the question "Where do you think those who developed and used transmission models work?" either "Prior to" or "During" the COVID-19 pandemic for both the online panel and social media samples. Percentages are taken with respect to the sample size of the platform (504 for the online panel and 202 for social media). As this question was open-ended, participants often provided more than one answer and so percentages do not represent mutually exclusive responses.*

| | **Online panel** | | **Social media** | |
|---|---|---|---|---|
| | **Prior to the COVID-19 pandemic** | **During the COVID-19 pandemic** | **Prior to the COVID-19 pandemic** | **During the COVID-19 pandemic** |
| **Advisory roles**<br><br>**[SAGE specifically]** | 2 (0%)<br><br>[0 (0%)] | 19 (4%)<br><br>[11 (2%)] | 0 (0%)<br><br>[0 (0%)] | 15 (7%)<br><br>[6 (3%)] |
| **Government** | 91 (18%) | 148 (29%) | 36 (18%) | 44 (22%) |
| **Healthcare services**<br><br>**[NHS specifically]** | 40 (8%)<br><br>[13 (3%)] | 73 (14%)<br><br>[32 (6%)] | 24 (12%)<br><br>[11 (5%)] | 22 (11%)<br><br>[10 (5%)] |
| **Media** | 10 (2%) | 8 (2%) | 2 (1%) | 6 (3%) |
| **Pharmaceutical industry** | 20 (4%) | 25 (5%) | 5 (2%) | 7 (3%) |
| **Public health bodies** | 41 (8%) | 59 (12%) | 56 (28%) | 57 (28%) |
| **Research (Academia)** | 166 (33%) | 183 (36%) | 141 (70%) | 148 (73%) |
| **Research (Unspecified affiliation)** | 176 (35%) | 153 (30%) | 36 (18%) | 32 (16%) |
| **Unsure** | 38 (8%) | 24 (5%) | 2 (1%) | 1 (0%) |
| **Other** | 115 (23%) | 102 (20%) | 26 (13%) | 38 (19%) |

***Supplementary Table 25:*** *Differences between where respondents thought transmission modellers worked across time periods within samples, according to Wilcoxon signed rank tests.* $p - values < 0.05$ *are considered statistically significant.*

| | | Number of respondents (n (%)) | | Wilcoxon signed rank test statistic | $n$ | $p - value$ |
|---|---|---|---|---|---|---|
| | | **Prior to the COVID-19 pandemic** | **During the COVID-19 pandemic** | | | |
| **Online panel** | **Advisory roles**<br><br>**[SAGE specifically]** | 2 (0%)<br><br>[0 (0%)] | 19 (4%)<br><br>[11 (2%)] | 22<br><br>0 | 504 | <0.001<br><br>[0.001] |
| | **Government** | 91 (18%) | 148 (29%) | 1196 | | <0.001 |
| | **Healthcare services**<br><br>**[NHS specifically]** | 40 (8%)<br><br>[13 (3%)] | 73 (14%)<br><br>[32 (6%)] | 313.5<br><br>119 | | <0.001<br><br>[0.001] |
| | **Media** | 10 (2%) | 8 (2%) | 7.5 | | 0.424 |
| | **Pharmaceutical industry** | 20 (4%) | 25 (5%) | 88 | | 0.284 |
| | **Public health bodies** | 41 (8%) | 59 (12%) | 367.5 | | 0.010 |
| | **Research (Academia)** | 166 (33%) | 183 (36%) | 630 | | 0.027 |
| | **Research (Unspecified affiliation)** | 176 (35%) | 153 (30%) | 3050 | | 0.021 |
| | **Unsure** | 38 (8%) | 24 (5%) | 304.5 | | 0.008 |
| | **Other** | 115 (23%) | 102 (20%) | 2464.5 | | 0.145 |
| **Social media** | **Advisory roles**<br><br>**[SAGE specifically]** | 0 (0%)<br><br>[0 (0%)] | 15 (7%)<br><br>[6 (3%)] | 0<br><br>0 | 202 | <0.001<br><br>[0.020] |
| | **Government** | 38 (19%) | 44 (22%) | 405 | | 0.230 |
| | **Healthcare services**<br><br>**[NHS specifically]** | 24 (12%)<br><br>[11 (5%)] | 22 (11%)<br><br>[10 (5%)] | 138<br><br>25 | | 0.683<br><br>[0.790] |
| | **Media** | 2 (1%) | 6 (3%) | 0 | | 0.072 |
| | **Pharmaceutical industry** | 5 (2%) | 7 (3%) | 22 | | 0.565 |
| | **Public health bodies** | 56 (28%) | 57 (28%) | 380 | | 0.879 |
| | **Research (Academia)** | 141 (70%) | 148 (73%) | 320 | | 0.266 |
| | **Research (Unspecified affiliation)** | 36 (18%) | 32 (16%) | 126 | | 0.383 |
| | **Unsure** | 2 (1%) | 1 (0%) | 1 | | 1.000 |
| | **Other** | 26 (13%) | 38 (19%) | 399.5 | | 0.078 |

***Supplementary Table 26:*** *Differences between where respondents thought transmission modellers worked within time periods, according to Chi-squared tests.* $p - values < 0.05$ *are considered statistically significant.*

| | | Number of respondents (n (%)) | | $\chi^2$ | $n$ | $p - value$ |
|---|---|---|---|---|---|---|
| | | Online panel | Social media | | | |
| **Prior to the COVID-19 pandemic** | **Advisory roles** | 2 (0%) | 0 (0%) | 0.01 | 706 | 0.910 |
| | **[SAGE specifically]** | [0 (0%)] | [0 (0%)] | [NA] | | [NA] |
| | **Government** | 91 (18%) | 38 (19%) | 0.02 | | 0.899 |
| | **Healthcare services** | 40 (8%) | 24 (12%) | 2.26 | | 0.132 |
| | **[NHS specifically]** | [13 (3%)] | [11 (5%)] | [2.88] | | [0.095] |
| | **Media** | 10 (2%) | 2 (1%) | 0.36 | | 0.548 |
| | **Pharmaceutical industry** | 20 (4%) | 5 (2%) | 0.55 | | 0.456 |
| | **Public health bodies** | 41 (8%) | 56 (28%) | 45.05 | | <0.001 |
| | **Research (Academia)** | 166 (33%) | 141 (70%) | 78.25 | | <0.001 |
| | **Research (Unspecified affiliation)** | 176 (35%) | 36 (18%) | 19.26 | | <0.001 |
| | **Unsure** | 38 (8%) | 2 (1%) | 10.38 | | 0.001 |
| | **Other** | 115 (23%) | 26 (13%) | 8.31 | | 0.004 |
| **During the COVID-19 pandemic** | **Advisory roles** | 19 (4%) | 15 (7%) | 3.44 | | 0.063 |
| | **[SAGE specifically]** | [11 (2%)] | [6 (3%)] | [0.12] | | [0.730] |
| | **Government** | 148 (29%) | 44 (22%) | 3.81 | | 0.051 |
| | **Healthcare services** | 73 (14%) | 22 (11%) | 1.31 | | 0.253 |
| | **[NHS specifically]** | [32 (6%)] | [10 (5%)] | [0.29] | | [0.593] |
| | **Media** | 8 (2%) | 6 (3%) | 0.80 | | 0.372 |
| | **Pharmaceutical industry** | 25 (5%) | 7 (3%) | 0.44 | | 0.507 |
| | **Public health bodies** | 59 (12%) | 57 (28%) | 27.44 | | <0.001 |
| | **Research (Academia)** | 183 (36%) | 148 (73%) | 77.62 | | <0.001 |
| | **Research (Unspecified affiliation)** | 153 (30%) | 32 (16%) | 14.97 | | <0.001 |
| | **Unsure** | 24 (5%) | 1 (0%) | 6.49 | | 0.011 |
| | **Other** | 102 (20%) | 38 (19%) | 0.11 | | 0.745 |

*How would you describe a transmission model?*

Descriptions of transmission models varied widely both within and across samples. Approximately 49% of the responses from online panel were deemed as relevant to the context of the research as defined in Methods in the main text, compared to 84% of social media respondents (Supplementary Figure 16; chi-squared test: $\chi^2 = 72.24$; $n = 706$; $p-value < 0.001$). In general, social media participants were more likely to use technical language such as "confidence interval" or to refer directly to mathematical model structures, such as "Susceptible, Infected, Recovered" than their online panel counterparts, highlighting potential differences in the experience and backgrounds of participants across samples. Nonetheless, the more relevant explanations of individuals were clear, regardless of sample, including:

"Theories that explain how diseases can be transmitted." (online panel respondent)

and

"A mathematical model which replicates the true transmission of the virus in the real world." (social media respondent).

The latter also highlights common occurrence across responses: the use of the term "the virus", or more generally, direct reference to COVID-19, despite this not being mentioned in the survey title or question. There were other interesting and common occurrences across both samples. For example, the "R number" was explicitly mentioned on multiple occasions, perhaps highlighting the continued use of this term at government COVID-19 press conferences and within the media more widely. Many participants also made use of the words akin to "graphic" in their model description, which could similarly be explained by considering that this was a common method of sharing the results of modelling with the public. The word "predict" or "prediction" was also commonly used by participants, which is particularly interesting given that experts have tried to emphasise the difference between scenario-based and predictive models.[3,5]

Approximately 20% of responses from the online panel were related to communication, for example: "A means to communicate information to someone, people or system.", compared to 3% of responses from social media. However, recall that social media respondents were likely to have accessed the survey directly from an account related to public health or mathematical modelling (Supplementary Table 1), and so the topic of the survey is likely to appear less "random" to this group, compared to the online panel respondents who saw this survey without any context. The other most common topics among online panel respondents included transmission modelling in the context of transmitting information (e.g., "A model whereby a message is sent from a source to a transmitter where they are then received and read.") and the sharing of information (e.g., "A guide to on how to disseminate information."). Social media respondents were borderline more likely to give an "opinion" as an answer, such as "production of false statistics" or "guess work erring massively over-cautious", than online panel respondents (3% and 1%, respectively; chi-squared test: $\chi^2 = 3.46$; $n = 706$; $p-value = 0.063$).

Most respondents from both samples selected being "moderately" or "very" confident in their model description (65% and 60%, respectively; chi-squared test: $\chi^2 = 1.34$; $n = 706$; $p-value = 0.247$), with "moderately confident" accounting for a majority of responses overall (Table 1). In both samples, being male was significantly associated with a higher level of confidence compared to being female, while within the online panel sample only the older age groups were associated with increased levels of confidence compared to the youngest age group (Supplementary Table 28).

A different and complex picture of confidence levels emerged when data were stratified by the relevance of the model description (Supplementary Figure 16). Although those with “more relevant” descriptions continued to primarily state being “Moderately confident” in their response, more respondents with less relevant model descriptions fell into one of the extremist categories of “Very” or “Not at all” confident (Supplementary Table 27). Among the “very confident” respondents, social media respondents were significantly more likely to have written a “more relevant” response than their online panel counterparts (81% and 29%, respectively; chi-squared test: $\chi^2 = 13.43$; $n = 706$; $p - value < 0.001$).

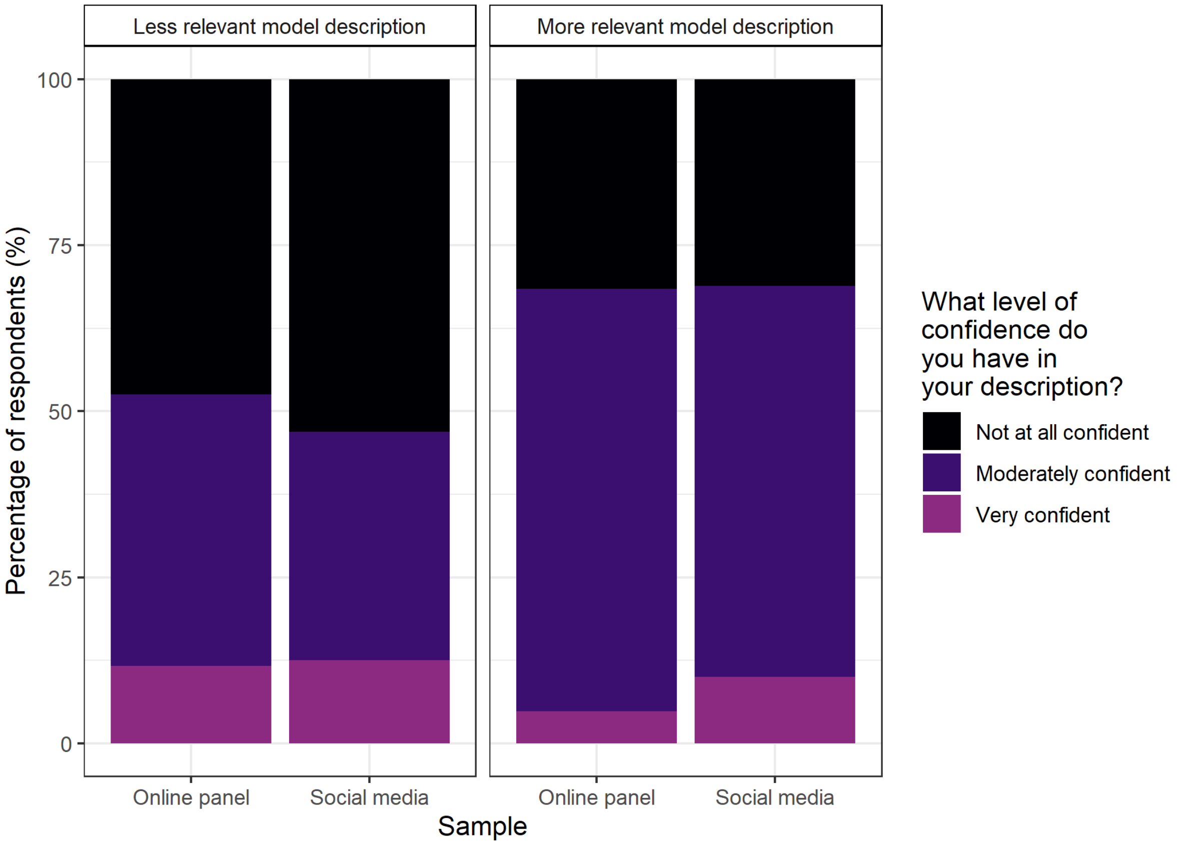

***Supplementary Figure 16:*** *The percentage of respondents from online panel and social media samples according to relevance of and confidence in answer provided to the question “How would you describe a transmission model?“. Underlying data presented in Supplementary Table 27.*

***Supplementary Table 27:*** *Number and percentage of respondents under each of the three confidence levels depending on the relevance of their description and their sample. Percentages calculated with respect to each column.*

| **What level of confidence do you have in your description?** | **Description less relevant** | | **Description more relevant** | |
|---|---|---|---|---|
| | **Online panel respondents** | **Social media respondents** | **Online panel respondents** | **Social media respondents** |
| **Not at all confident** | 122 (48%) | 17 (53%) | 78 (32%) | 53 (31%) |
| **Moderately confident** | 105 (41%) | 11 (34%) | 157 (64%) | 100 (59%) |
| **Very confident** | 30 (12%) | 4 (13%) | 12 (5%) | 17 (10%) |
| **Total** | 257 (51%) | 32 (16%) | 247 (49%) | 170 (84%) |

***Supplementary Table 28:*** *Coefficient estimates, standard errors and p-values for the linear regression models with gender and age group as predictors of level of confidence within each model for each sample. The three levels of confidence provided in the multiple-choice options, "Not at all confident", "Moderately confident" and "Very confident", were enumerated as -1, 0, 1, respectively.* $p - values < 0.05$ *are considered statistically significant.*

| **Sample** | **Response** | **Variable** | **Interpretation** | **Estimate** | **Standard Error** | **p-value** | **Chi-squared test** | |
|---|---|---|---|---|---|---|---|---|
| | | | | | | | $\chi^2$ | **p-value** |
| Online panel | What level of confidence do you have in your description? | Intercept | Baseline* | -0.68 | 0.08 | <0.001 | | |
| | | Age (years) | 26 – 35 | 0.15 | 0.10 | 0.107 | 7.29 (df=5) | 0.001 |
| | | | 36 – 45 | 0.37 | 0.09 | <0.001 | | |
| | | | 46 – 55 | 0.35 | 0.10 | <0.001 | | |
| | | | 56 – 65 | 0.28 | 0.09 | 0.002 | | |
| | | | 66+ | 0.32 | 0.12 | 0.009 | | |
| | | Gender | Male | 0.23 | 0.05 | <0.001 | 6.92 (df=3) | <0.001 |
| | | | Non-binary | 0.52 | 0.60 | 0.389 | | |
| | | | Prefer not to say | 0.41 | 0.35 | 0.245 | | |
| Social media | What level of confidence do you have in your description? | Intercept | Baseline* | -0.24 | 0.31 | 0.445 | | |
| | | Age (years) | 26 – 35 | 0.25 | 0.36 | 0.492 | 2.58 (df=5) | 0.225 |
| | | | 36 – 45 | -0.20 | 0.33 | 0.536 | | |
| | | | 46 – 55 | -0.14 | 0.31 | 0.656 | | |
| | | | 56 – 65 | -0.19 | 0.31 | 0.534 | | |
| | | | 66+ | -0.24 | 0.33 | 0.459 | | |
| | | Gender | Male | 0.32 | 0.09 | <0.001 | 5.10 (df=2) | 0.001 |
| | | | Prefer not to say | 0.43 | 0.36 | 0.232 | | |

*Baseline comprised of "Female" and "18 – 25" years.
